# The impact of digital contact tracing on the SARS-CoV-2 pandemic - a comprehensive modelling study

**DOI:** 10.1101/2020.09.13.20192682

**Authors:** Tina R Pollmann, Julia Pollmann, Christoph Wiesinger, Christian Haack, Lolian Shtembari, Andrea Turcati, Birgit Neumair, Stephan Meighen-Berger, Giovanni Zattera, Matthias Neumair, Uljana Apel, Augustine Okolie, Johannes Müller, Stefan Schönert, Elisa Resconi

## Abstract

Contact tracing is one of several strategies employed in many countries to curb the spread of SARS-CoV-2. Digital contact tracing (DCT) uses tools such as cell-phone applications to improve tracing speed and reach. We model the impact of DCT on the spread of the virus for a large epidemiological parameter space consistent with current literature on SARS-CoV-2. We also model DCT in combination with random testing (RT) and social distancing (SD).

Modelling is done with two independently developed individual-based (stochastic) models that use the Monte Carlo technique, benchmarked against each other and against two types of deterministic models.

For current best estimates of the number of asymptomatic SARS-CoV-2 carriers (approximately 40%), their contagiousness (similar to that of symptomatic carriers), the reproductive number before interventions (*R*_0_ at least 3) we find that DCT must be combined with other interventions such as SD and/or RT to push the reproductive number below one. At least 60% of the population would have to use the DCT system for its effect to become significant. On its own, DCT cannot bring the reproductive number below 1 unless nearly the entire population uses the DCT system and follows quarantining and testing protocols strictly. For lower uptake of the DCT system, DCT still reduces the number of people that become infected.

When DCT is deployed in a population with an ongoing outbreak where 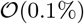 of the population have already been infected, the gains of the DCT intervention come at the cost of requiring up to 15% of the population to be quarantined (in response to being traced) on average each day for the duration of the epidemic, even when there is sufficient testing capability to test every traced person.

## 1 Introduction

Tracing and isolation of people who were in contact with an infectious person (contact tracing) can be used to control the spread of communicable diseases [1, 2]. In the traditional understanding of contact tracing (CT), public health employees interview known carriers (index cases) of the disease and then track down people who had the type of close contact with the index case necessary to transmit the disease. Contacts are then diagnosed and isolated. This implementation is only suited for infections that spread relatively slowly, and where cases can be easily diagnosed [3]. SARS-CoV-2, with its unspecific symptoms, high number of asymptomatic carriers, and incubation times as short as a day, does not fit this mold. Recent studies indicate that an outbreak of SARS-CoV-2 could be controlled using fast and efficient digital contact tracing (DCT) [4, 5]. DCT systems using cellphone applications based on Bluetooth proximity measurements are currently being developed and/or deployed in many countries [6, 7, 8, 9, 10, 11, 12, 13, 14, 15]. Predicting the effect of DCT on an outbreak is challenging, especially since the epidemiological parameters that describe the outbreak dynamics are currently still uncertain. Almost no practical experience for DCT is available. Partly automated CT systems were in use during the Ebola outbreak 2014-2016 [16] and it turned out that the technical difficulties that come with that approach must not be underestimated.

Simulation studies focusing on classical CT for COVID-19 consistently indicate that around 70% of the contacts need to be traced, and that the tracing delay has to be as short as 1 day [5, 17]. For these reasons, [17] doubt that COVID-19 can be controlled by traditional CT in practice. Ferretti et al. [4] however point out that DCT could significantly reduce tracing delays, so that the outbreak could be controlled for tracing probabilities much smaller than 70%. Meanwhile, several other simulation studies indicate that a high tracing probability and a combination of fast CT and testing is required to control SARS-CoV-2 [16]. However, it also became clear that not only the reduction of the reproduction number, or the final size of the epidemic needs attention, but also the number of persons that go to quarantine are of importance [18]: A naive application of DCT leads to a situation that resembles a lock-down as a large fraction of the population is quarantined. As we also find in the present study, an appropriate choice of the tracing and testing protocol is central.

We developed individual-based models with the Monte Carlo (MC) simulation technique, flanked and cross-checked by deterministic models, to evaluate the dynamics of a COVID-19 outbreak under different intervention protocols, focusing on DCT and DCT combined with random testing and social distancing. We determine not just the immediate effective reproductive number *R_e_*, but also the daily *R_e_*, the number of healthy people in quarantine, and the number of people infected, for up to a year of continuous interventions. The sensitivity of all outcomes to the reported ranges of values of the epidemiological parameters is studied in detail.

The goal of this paper is to study quantitatively under which conditions and to which degree DCT combined with fast testing and social distancing can replace rigorous shelter-in-place policies for keeping the effective reproduction rate *R_e_* ≤ 1.

## 2 Model inputs

Table 1 presents an overview of all model input parameters and the values considered for them. They will be discussed in detail in the following subsections.

Many model inputs that are properly described by a distribution rather than just a mean value are modelled using a shifted Gamma distribution, defined as

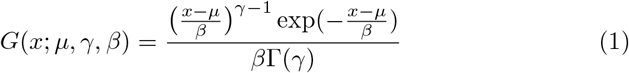

where *x* ≥ *μ*. As this distribution is flexible, we also use a discretized version (*n* ∈ ℕ_0_)

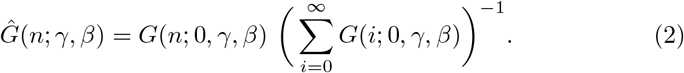

### 2.1 Social contact structure

Our deterministic models describe contacts between individuals in a population by the usual mass action law. The stochastic models allow for a more detailed investigation. Particularly, two different strategies are considered: For the first, in the following referred to as *homogeneous population*, we choose the contacts for each person randomly out of the entire population. The probability for a person to have *n* unique contacts close enough to transmit a respiratory virus on a given day is taken from the empirical distributions reported in [19]. These are well described by

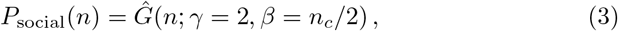

where *n_c_* is the mean number of contacts per day. We also consider a more realistic contact pattern, in the following referred to as *social graph population*. The population is described by a social graph, were each individual is represented by a node and contacts are represented by edges. Each individual is given a fixed set of contact persons for the entire simulation. We employ a modified version of the *Lancichinetti–Fortunato–Radicchi* benchmark graph (LFR) [20] as shown in [21] (therein referred to as LFR-BA). In this model, the population is divided into communities with sizes distributed according to a power-law distribution. Node edges are constructed according to the linear preferential attachment model [22] under the constraint that an average fraction of (1 − *μ*) of the edges of each node connect nodes within the same community. A graph constructed in this manner results in a power-law probability distribution with index *a* = 2 for the node degree *n* [22]:

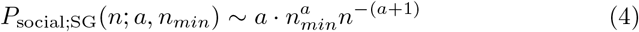

where *n_min_* is the minimum node degree. Here, the mean number of contacts per day is given by 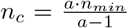. We assume that each edge is active once per day, so that the node degree corresponds to the number of contacts per day.

### 2.2 Epidemiological parameters

Fig. 1 schematically shows the probability distributions for symptom onset, transmission, and true positive test results. The parameters are explained in the following paragraphs.

**Figure 1:**
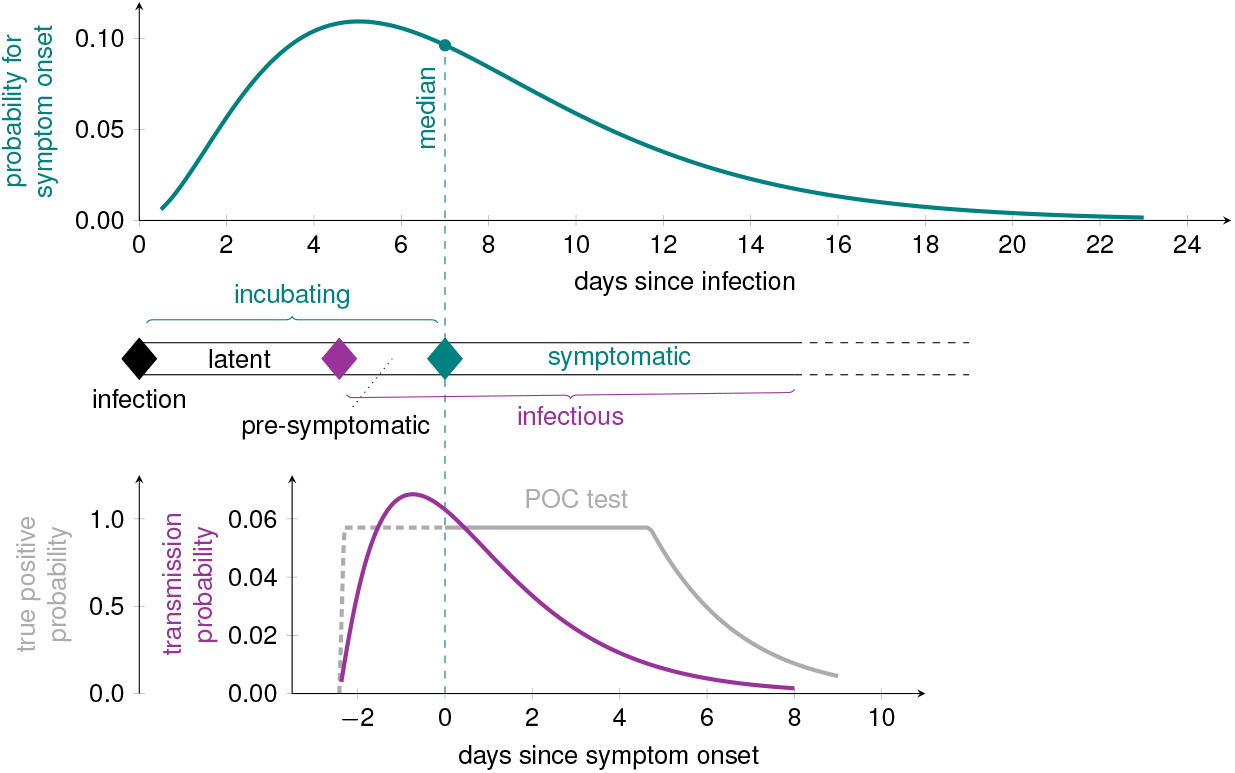
Course of the disease with probability distribution for the incubation period *T*_Inc_ (top) [23] and the fully correlated probability density function for the contagiousness *T*_con_ (bottom) [24]. The dotted vertical lines corresponds to the median of *T*_Inc_. The probability for a true{positive point-of-care (POC) test is displayed on the bottom left (grey line). The diamonds correspond to exposure (contact, black), end of latency/begin of contagious period (magenta) and symptom onset (teal).

#### Incubation period P(*T*_Inc_)

The distribution of incubation periods is taken as

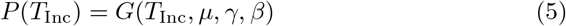

(Fig. 1 upper curve) with shape parameters chosen to match the curve reported in [23], which has a median and mean incubation period of 7 days and 7.44 days, respectively, with a range from 0-23 days. The authors included a relatively large group of patients (n=587) with a wide age (0-90 years) and symptom (asymptomatic to severe) range. Other studies, albeit with smaller number of patients and with a bias towards more severe symptoms, have reported lower medians [25, 26]. Therefore, in addition to the parameters matching [23] (printed in black in Table 1) we also model a curve with shorter median and mean incubation period of 3.6 days and 4 days (printed in gray in Table 1).

#### Latent period *T*_lat_

The latent period for SARS-CoV-2 is shorter than the incubation period, meaning pre-symptomatic transmission can occur [1][27]. The latent period is difficult to determine empirically, as it requires exact information about the time of exposure and contagiousness. As, to our knowledge, no reliable, large-scale studies have been published on the latent period of SARS-CoV-2 so far, we use the measured contagiousness relative to the incubation time as an auxiliary mean to infer latency,

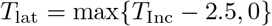

where the value of 2.5 days comes from the transmission probability curve discussed in the next paragraph.

#### The transmission probability curve *P*_trans_(*τ*)

The transmission probability is the probability that a contagious person infects someone they have contact with. This probability is often given as an average “infectivity per day”, *β_i_*, even though it changes significantly as a function of the time since infection *τ*. The infectivity was measured as a function of the time since onset of symptoms by He et al. [24]. They find that carriers become contagious approximately 2.5 days before the onset of symptoms, and that approximately 44% of transmissions occur during this pre-symptomatic phase.

We take as the contagious period *T*_con_ the time from the end of the latent period until 99% of the cumulative transmission probability is reached; a person is considered to be recovered afterwards. Therewith, the transmission probability as a function of time since infection is given by a scaled and truncated Gamma distribution (Fig. 1 lower curve). After infection, an individual has a (random) latent period *T*_lat_, during which the transmission probability is zero. Afterward, for *τ* = *T*_lat_ + *t* (and *t* ≥ 0), we have

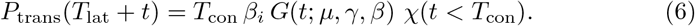

where *χ* is 1 if the condition in the argument is met, and 0 otherwise.

The shape parameters shown in black in Table 1 are our defaults, taken to match the curve in [24]. Pre-symptomatic infectivity is strongly debated. Therefore, we model a second shape where only 18% of the transmission occurs during the pre-symptomatic phase (shape parameter values printed in gray).

The course of the disease for asymptomatic carriers is the same as that for symptomatic carriers as shown in Figure 1, and the incubation time is then the time when symptoms would have started.

#### Fraction of asymptomatic cases 1 − *α*

The proportion of asymptomatic carriers (1 − *α*) described in the literature ranges from ~ 4% [28] to ~ 40%[29] of all cases. Initial reports for (1 − *α*) derived from testing of specific cohorts (cruise ship, returning travellers) ranged from 17-31% [27, 30, 31], however larger studies suggest even higher numbers. Analysis of the mass screening of the full population of the municipality of Vo’, Italy [29] report that 41.1% of the confirmed SARS-CoV-2 infections were asymptomatic (as defined by the absence of fever and/or cough). Ferretti et al. [4] analyzed 40 selected transmission pairs and also derived a value of 40% for the proportion of asymptomatic infected individuals. We model several different values for *α* to cover the reported ranges.

Reduced asymptomatic transmission probability *η_as_*.

While initial studies assume that asymptotic cases are less contagious [32, 4], newer reports indicate that the viral load of asymptomatic cases is similar to symptomatic cases, which suggests similar contagiousness [33, 34, 35, 36, 37, 38]. We nevertheless introduce the parameter *η_as_* ∈ [0,1], which scales the transmission probability for pre- and asymptomatic cases, and vary that parameter to explore its effects. We note that when we model with *η_as_* < 1, we apply the scaling to both pre- and asymptomatic phases; since there most likely is no difference in viral load, if asymptomatic transmission is suppressed, this is likely due to circumstantial factors, like lack of coughing, which apply to the pre-symptomatic phase as well.

The basic reproductive number *R*_0_.

If the contagiousness is independent of the symptom status, i.e. *η_as_* =1, the reproductive number is given by the number of contacts during the contagious period, *n_c_T*_con_, times the average probability to transmit the infection in one contact, *β_i_*. However, we need to distinguish between symptomatic, pre-, and a-symptomatic cases in order to allow for *η_as_* < 1. We find

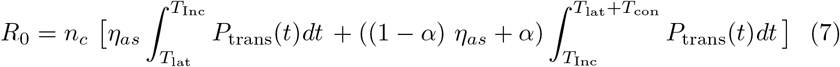

Note that – though *P*_trans_ is a random function – the random part of the function is a pure translational offset (the latency period), s.t. the integral is deterministic.

*R*_0_ is expected to be different in different populations because *n_c_* is different by factors of up to 4 just within the populations of different European countries. Values of *R*_0_ ranging from 1.4 to 6.5 have been reported [43], though it is not always clear whether or not the reported *R*_0_ is for the case where symptomatic individuals are quarantined. Furthermore, *R*_0_ is usually not corrected for the contact rate in the population where it is studied. We consider *R*_0_ the reproductive number without any form of interventions. We take *R*_0_ of 3 (approximately the median reported in [43]) as our default, but also run the models for *R*_0_ of 2 and 4.

Following [44], we introduce the random reproduction number for an individual *R*_0_,*i*. Then, of course, *R*_0_ is the expectation value over the *R*_0_,*i*. For simplicity (and since two of our models are not based on continuous time but on discrete time/days), we state the time-discrete formula for *P*(*R*_0_,*i* = *n*). Assume that an individual did infect *n* persons. These *n* persons can be arbitrarily distributed over the contagious period. In a slight abuse of notation, let *T_c_* denote the number of days that a person is contagious for. Furthermore, let 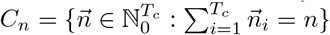 be all possible ways to distribute the *n* infectees to *T_c_* days. Then, for a homogeneous population,

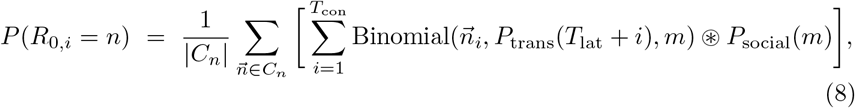

where the convolution ⊛ is over the parameter *m*. This distribution is shown for some parameter combinations in Fig. 2.

**Figure 2:**
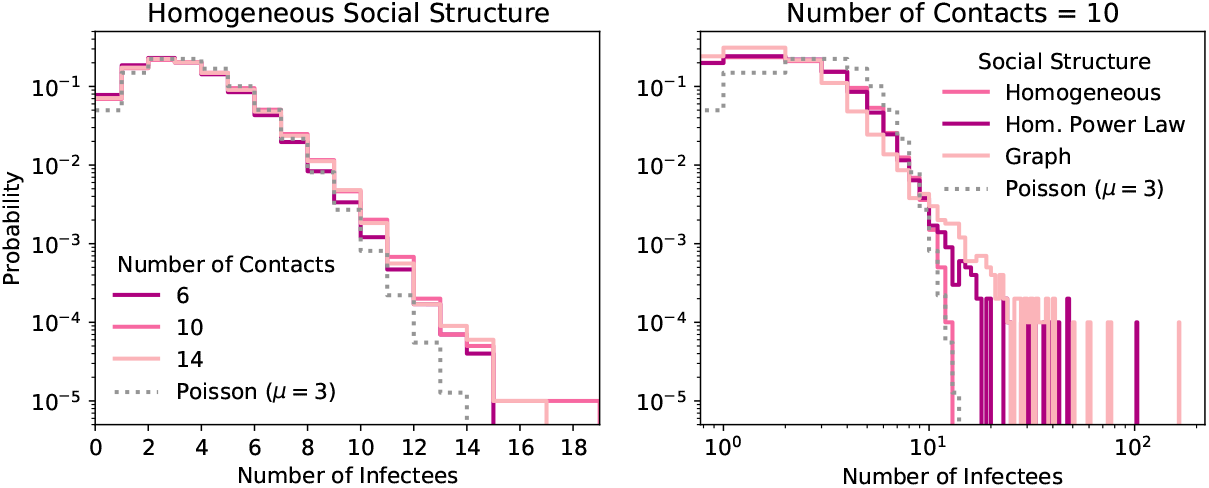
The distribution of the number of people a carrier infects (Eq. (8)) for 3 combinations of *n_c_* (the mean number of contacts per day) and *β_i_* (the average transmission probability per day) that result in *R*_0_ =3 (see Tab. 1) (left) and for a fixed *n_c_* but different social structures (right). Default values for the infection probability curve are used, *η_as_* = 1, and no interventions are applied. Compared to a Poisson distribution with mean of 3, the distribution is over-dispersed.

### 2.3 Intervention protocols

The interventions considered here are 1) DCT, 2) quarantining, 3) testing, and 4) social distancing. Reported symptomatic cases are quarantined starting right at the beginning of the epidemic. The remaining interventions are turned on once a fraction *f_i_* of the population has become exposed.

**(1) DCT**. We assume that a fraction *p*_app_ of the population uses the DCT system and that we can trace all contacts between users of this system with time delay *T*_delay_. In case that both infector and infectee have a DCT device, the probability for successful tracing is *η_DCT_*, while tracing always fails if either infector or infectee do not use the system. *η_DCT_* accounts for situations where cell phones run out of battery, are not with the owner at all times, Bluetooth is turned off, or where an alert is ignored. The DCTS system identifies contacts within the past Δ*T_trace_* days.

In the literature, the overall tracing probability across the population is often taken as 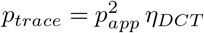. We note that this formula is not correct but becomes approximately right if 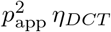 is small (see supplemental materials).

To become an index case for tracing, a person must be reported. We assume that from the group of symptomatic carriers, a fraction *f_m_* sees a doctor to get tested with a reliable laboratory test and is then reported. A fraction (1 − *α)* of cases will go unreported because they do not exhibit symptoms, unless they get tested due to being traced. A fraction *α*(1 − *f_m_)* of symptomatic cases will go unreported due to lack of access to medical tests. In the case where *η_as_* =1, *α* and *f_m_* are degenerate.

First order tracing refers to a protocol where contacts of an index case are traced. DCT also allows immediate tracing of contacts-of-contacts. We refer to this as second order tracing. If the Δ*T_trace_* is big enough, DCT will identify the infector. Second order tracing then can trace not just the people infected by an index case, but also the people who were infected by the same infector as the index case.

A DCT system will identify all contacts, regardless of their infection status. Since many models assume perfect accuracy in identifying only contacts that became infected, we run all parameters both with (closer to reality) and without (to be comparable to other models) tracing of the uninfected contacts.

All traced people immediately go into quarantine. This is necessary to suppress the pre- and asymptomatic transmission rates.

**(2) Quarantining**. Quarantining refers to any intervention that reduces the transmission probability significantly; this includes self-quarantine at home as well as being hospitalized. We assume that all reported symptomatic patients are immediately quarantined, regardless of any other interventions. This already reduces the reproductive number to

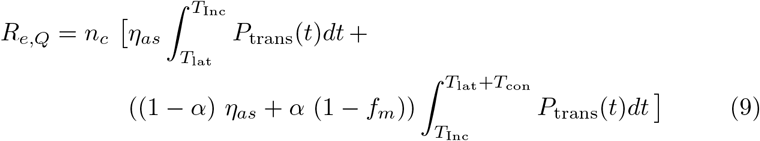

Fig. 3 shows *R_e,Q_* for combinations of *α*, *f_m_*, and *η_as_*. In Fig. 3 (top), *η_as_* =1, so only the product of *a* and *f_m_* is relevant, and results are shown for *R*_0_ =3 and for *R*_0_ =2. In Fig. 3 (bottom), *η_as_ <* 1, so both *R*_0_ and *R_e,Q_* depend on *α* and on *f_m_*. For each combination of *α* and *f_m_*, the transmission probability *β_i_* was adjusted to obtain *R*_0_ =3.

**Figure 3:**
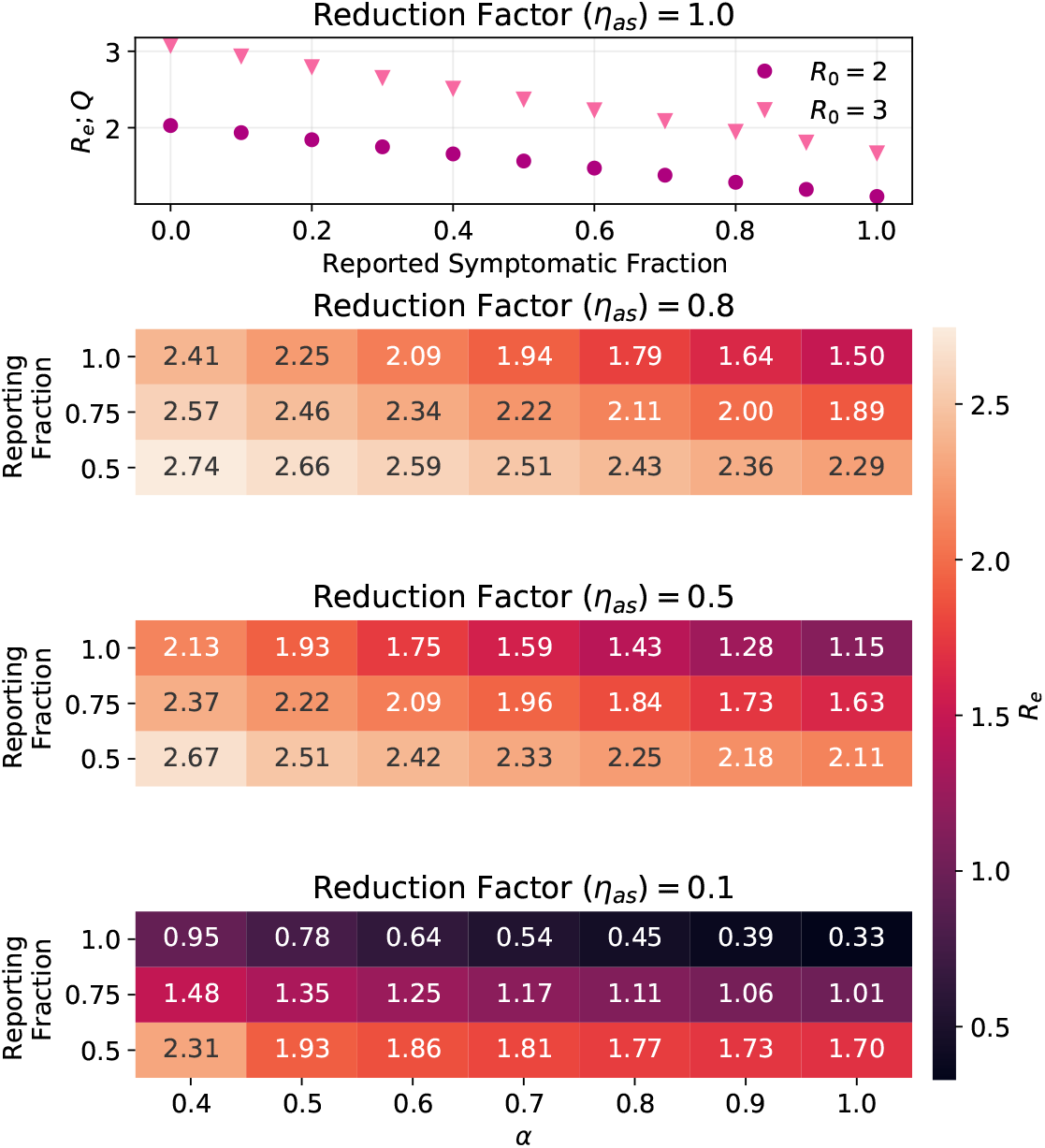
The effective reproductive number reached just from quarantining reported symptomatic carriers, *R*_e;Q_, is shown for four different values of *η_as_* (asymptomatic infectivity scaling) as calculated from Eq. (9). Top panel: In the case of *η_as_* =1, *R*_e;Q_ depends only on the product of *α* (symptomatic fraction) and *f_m_* (fraction reported and tested) and is shown for two values of *R*_0_. Lower three panels: For each combination of *η_as_* and *α*, the infection probability was adjusted to obtain *R*_0_ = 3.

In response to being reported or being traced, people are quarantined by default for 14 days. Symptomatic cases may leave quarantine 8 days after the symptoms start. Uninfected contacts can leave quarantine early following a testing protocol.

**(3) Testing**. We consider two types of tests. A reliable laboratory test for symptomatic carriers seeing a medical professional, and a fast point of care (POC) test that can be performed at home or at mobile testing stations. In either case, carriers only test positive while they have a high enough viral load. We assume the tests have *p*^true positive^ = 0.0 while the carrier is in the latent period, that is up to approximately 2.5 days before symptom onset. The viral load rises quickly after the end of the latent period. We further assume that the laboratory test then has a true positive rate of 100% until the carrier has recovered. The POC test on the other hand has 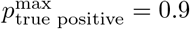 until approximately 5 days after symptom onset. After this time, the true positive rate falls at the same rate as the transmission probability curve; the true positive rate as a function of days since symptom onset is shown in Fig. 1 (bottom gray curve).

All people who are traced must be tested for two reasons: a) A positive test result is the only way for asymptomatic individuals to become index cases for tracing, and index cases are needed for tracing to be effective, and b) so uninfected traced people can be released from quarantine. Keeping all traced people in quarantine for the full quarantining duration means that a large fraction of the uninfected population may end up quarantined on any given day of the outbreak. We use the following release protocol: All traced people go into quarantine and get tested with a POC test. Regardless of the test result, everyone stays in quarantine, because the person may still be in the latent period. Those who tested negative on the first day are re-tested *δT_re-test_* days later. If both tests were negative, the person may leave quarantine, but is tested again after another *δT*_re-test_ days in case they were still in the latent period when the second test was done.

In addition to testing in response to being traced, we simulate the option of randomly testing a fraction *f*_RT_ of the population each day. This is done with a testing protocol assumed to have a negligible number of false positives.

**(4) Social Distancing**. Social distancing includes both a reduction of the total number of contacts per day *n_c_* to *n_c_* · *f*_SD_, and limiting the maximum number of contacts per day. In the absence of second-order effects, and if the upper limit does not change the distribution mean significantly, this reduces the reproductive number to

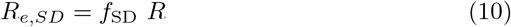

where *R* is the reproductive number without social distancing.

**Table 1:**
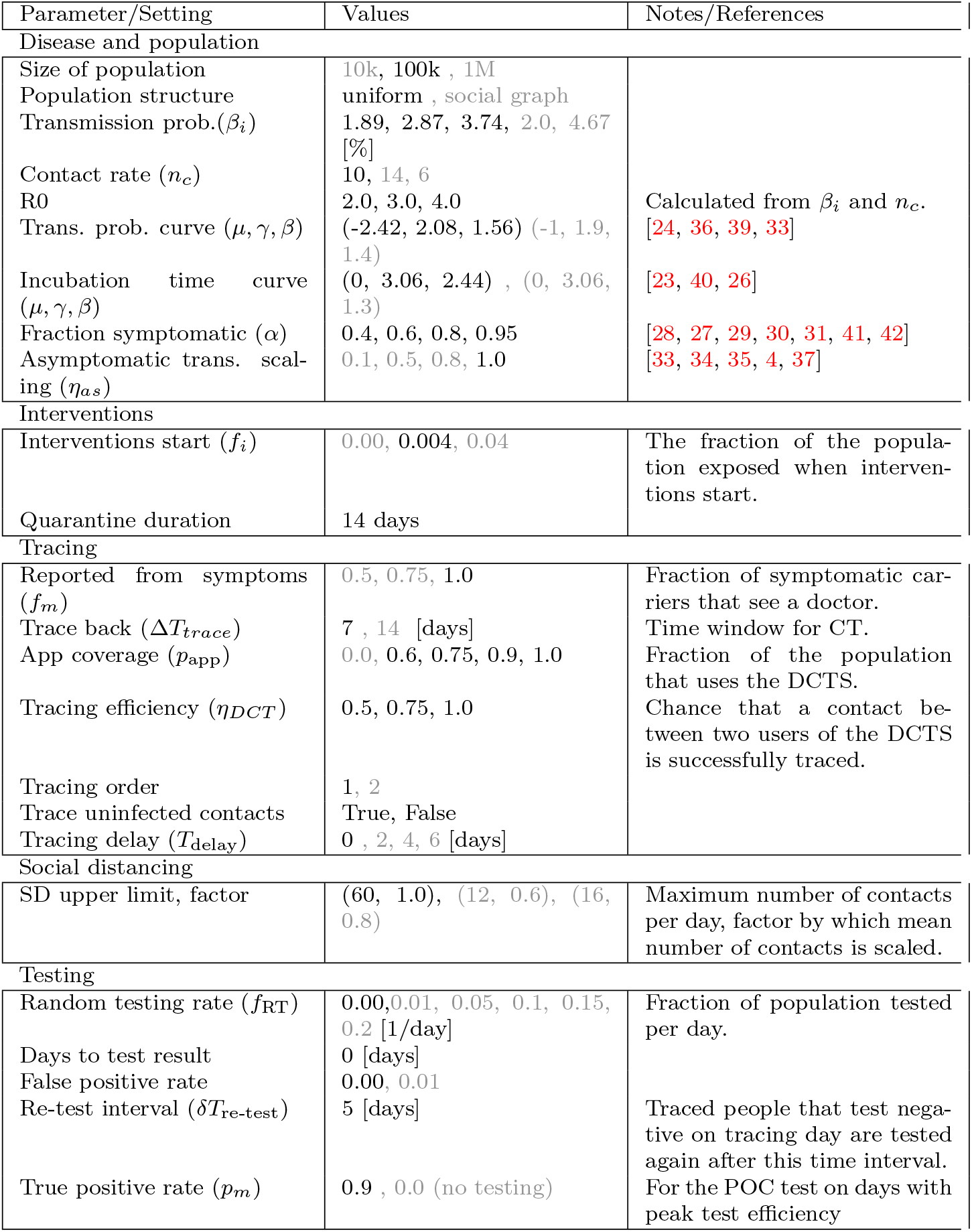
Key model input parameters and settings. The models are evaluated for all possible combinations of all parameter values shown in black. Parameter values in grey are used only with select other parameter combinations.

## 3 Models

Epidemiological modelling is a well established scientific discipline and different approaches, including contact tracing, are described in the rich literature [45, 46, 47, 48]. Epidemiological models that account for CT date back to the 1980s [49]. The main challenge to modelling a CT system is the individual-based character of CT, and the handling of the resulting stochastic dependencies between individuals. Individual-based simulation models [50] readily describe this process. For the scope of this paper, we developed two deterministic and two individual-based models. This redundant approach serves to cross-validate the results.

### 3.1 Deterministic models

The early phase of an outbreak can be quantitatively described with compartmental models based on ordinary differential equations (ODE) [51] or with age-since-infection models [52].

The deterministic models used here bridge the different scales utilizing the mathematical analysis of the underlying, microscopic stochastic branching process with contact tracing. The effect of contact tracing on the removal rates are determined. These effective removal rates are then used in the deterministic models. Our first deterministic, compartmental model explicitly predicts the status (exposed/infectious) for a newly infected person, when he/she will eventually be traced. Eventually traced and never-traced individuals go to different compartments. In that, the (exponentially distributed) waiting times can be readily adapted. Particularly, the model is close to standard SEIR-models (see Fig. 4), and is feasible to analytical analysis (Appendix A). In contrast, the second model, based on age since infection, does not explicitly formulate an exposed and an infectious period. The basic assumption is that the state of an individual is a function of his/her age of infection, that is the time that has passed since he or she became infected. The structure is less pronounced, but it is possible to use transition rates that rate more realistic (Appendix B). At the present time, the analytical treatment of the interdependence of contact tracing and correlations between infected individuals at the plateau phase of an epidemic is not well understood. Therefore, both models focus on the onset of the epidemics, where the reduction of susceptible by quarantine does not play a central role. The main outcome of these two models are the doubling time *T*_2_ and effective reproductive number *R_eff_* for interventions starting on the first day of the epidemic, though both models are able to predict in a heuristic way the total course of the epidemics.

**Figure 4:**
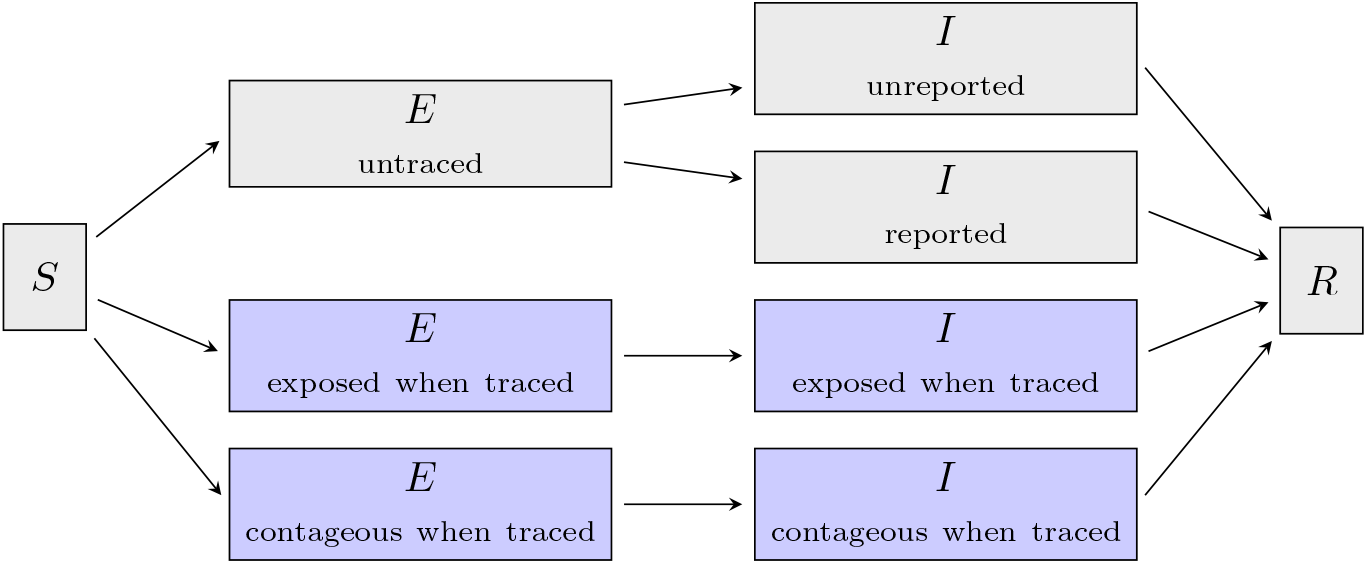
Simplified structure of the compartmental model. The model is based on a SEIR-type model from Ref. [53] and distinguishes between untraced (gray) and traced (blue) individuals. A detailed description of the model and its variables can be found in Appendix A.

### 3.2 Individual-Based Models

We developed two independent individual-based models (IBMs), which use the Monte Carlo (MC) technique to simulate social interactions, the progression of the viral disease, and interventions, at the level of individual people. The MC simulations proceed through the outbreak in steps of one day. Each day of the outbreak, every infected person not in quarantine meets a number of other people randomly drawn either from *P*_social_(*n*) (for a homogeneous population structure) or from the person’s social graph. The probability to infect each contact is given by *P*_trans_(*τ*). When a contact becomes infected, the incubation time is drawn from *P*(*T*_Inc_). The intervention protocols are implemented as described in Sect. 2.

Figure 5 shows a chain of infections from one of the simulation runs. Each box represents a person, and arrows between boxes represent infections and tracing.

**Figure 5:**
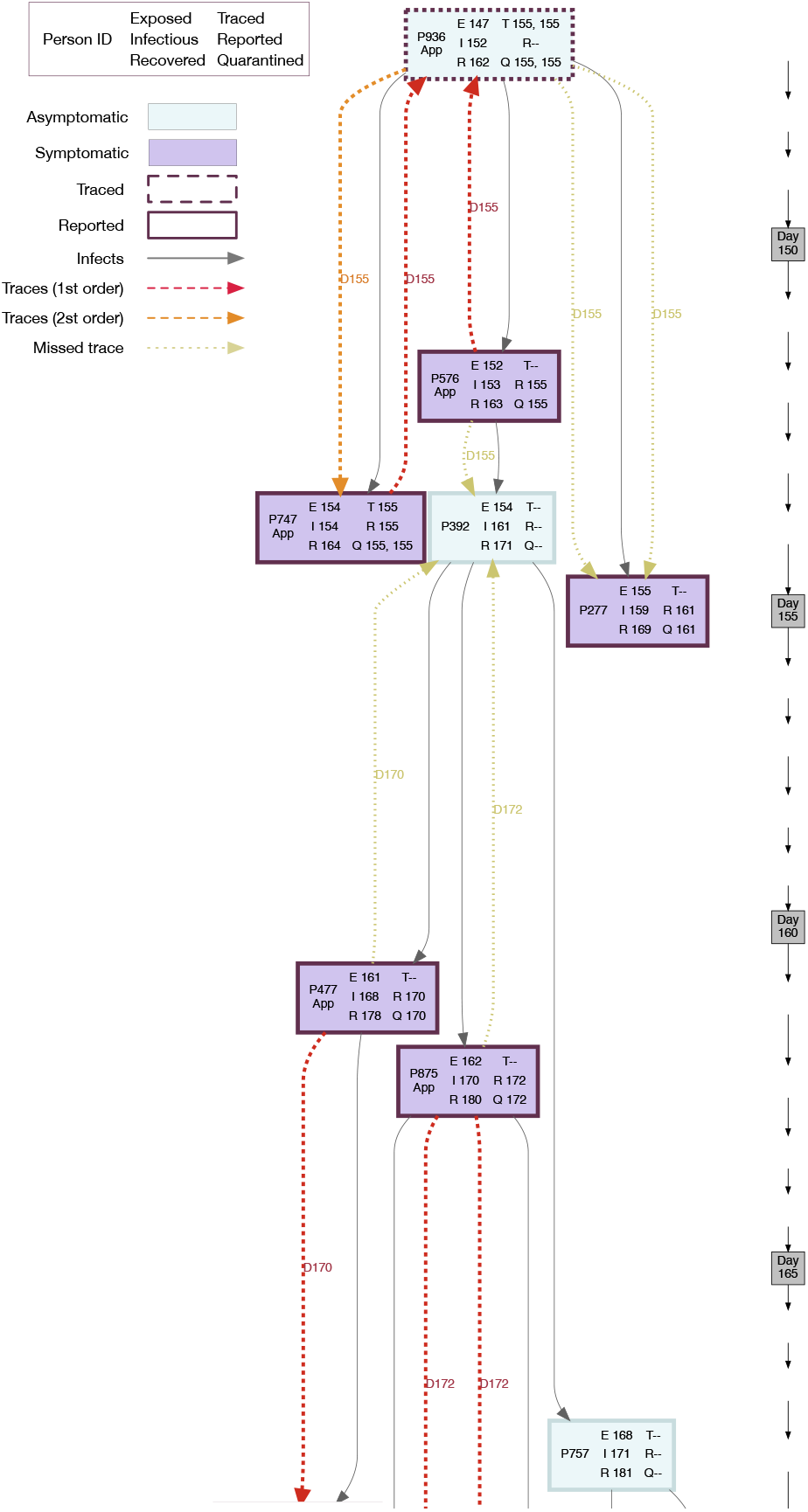
Spread and containment of SARS-CoV-2 using tracingand quarantine interventions in a Monte Carlo simulation including 1st; and 2nd order tracing. The downward arrows at the right indicate the time axis, with days in the simulated epidemic given in the grey boxes. Shown are the first 3 generations of infectees originating from simulated person P936. The input settings for the simulation were: *R*_0_ =2.0, Δ*T*_trace_ =7 days, *p*_app_ =0.6, *η_DCT_* =0.9, *α* =0.6. Each box represents a person. The leftmost column in each box gives the person ID and whether or not they use the DCT app. The middle column indicates the days when the person was exposed, became infectious, and recovered. The rightmost column indicates if and when the person was traced, reported, and quarantined.

In this example, P936 was exposed to the virus on day 147 of the simulated epidemic and had a latent period of 5 days, but never developed symptoms (light blue background) or tested positive and was therefore never reported (R–). He or she infected 3 others – P576 on day 152, P747 on day 154, and P277 on day 155. All three infectees developed symptoms (purple background). P576 saw a doctor on day 155, tested positive, and become reported. This triggered tracing of his infector, P936, and of the person he or she infected, P392. Tracing to P392 failed because this person does not use the app. Since P392 also does not develop symptoms, he or she never gets reported or quarantined and infects 3 others. The backward trace from P576 to P936 puts P936 in quarantine on day 155 and thus prevents him or her from infecting more people after this time. P936 does not tests positive (dashed outline of the box indicates the person was traced but never reported), so never becomes an index case him- or herself. However, since second order tracing is active in this simulation, the ‘siblings’ of P576 are identified. P747 is put in quarantine before he or she can infect anyone else, and tests positive the same day. This makes him or her an index case, so that the common infector, P936, is traced again. P277 does not use the app, therefore the trace fails. However, P277 happens to not meet many people on the first 2 days of infectiousness, then developed symptoms, saw a doctor, and was quarantined.

In this example, the chain of infections was stopped at P747 through second order tracing, the chain was stopped at P277 due to luck, but the chain could not be interrupted at P576 because the person he or she infected did not use the tracing app.

For a given set of input parameters, that is for a specific scenario, each run of the MC simulation represents one possible course of the epidemic. To find the most likely outcome for a scenario, the simulation is run 50 to 1000 times and the outcomes are averaged. As an example, Figs. 6 and 22 show the course of the epidemic for 20 MC runs each. The stochastic nature of the processes involved creates a spread in outcomes. Especially near the beginning of the epidemic where only few people are infected, statistical fluctuations cause large differences in the outbreak dynamics.

**Figure 6:**
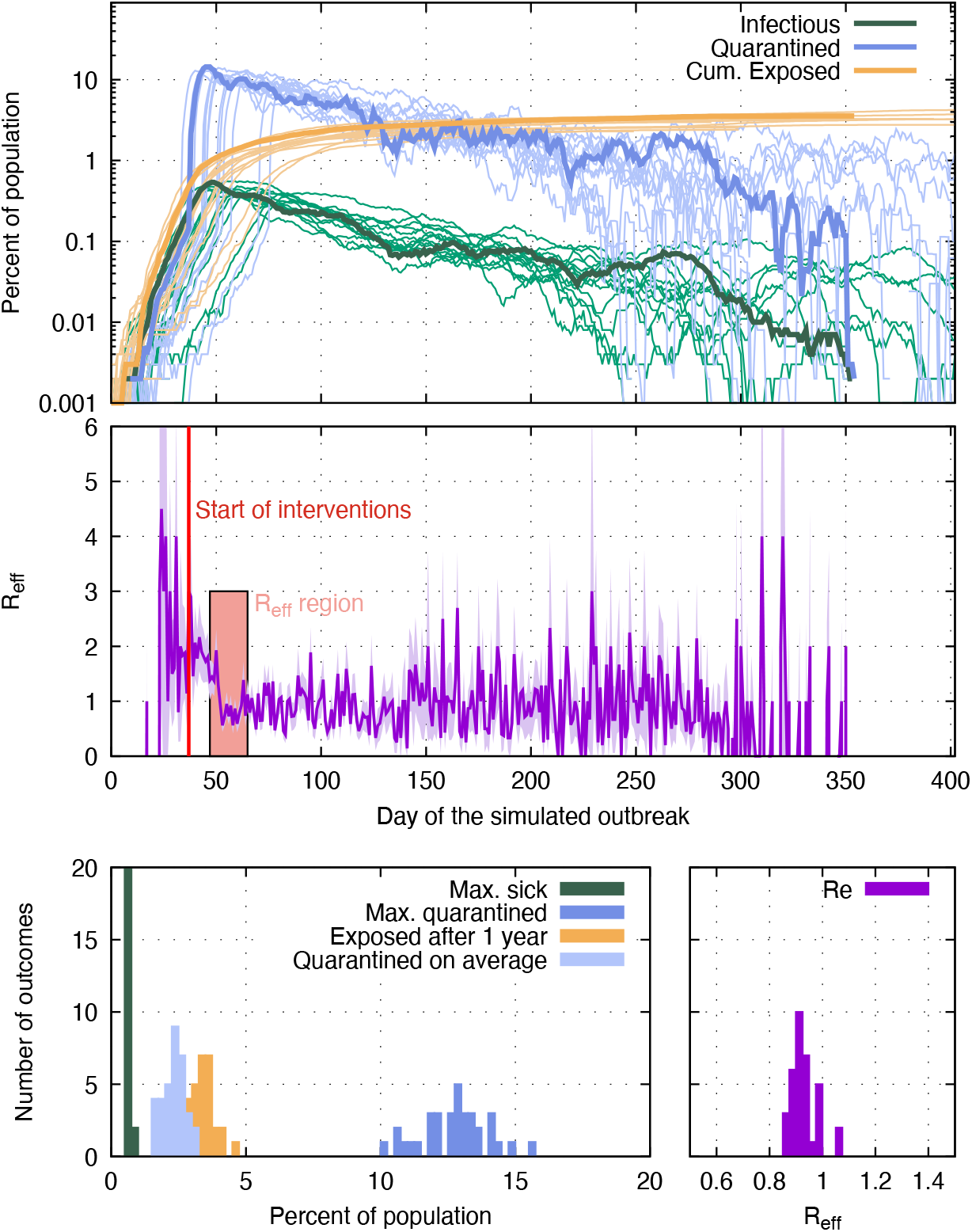
Stochastic variation in outbreak dynamics. The results are from 50 runs of the MC simulation; each run has the same input parameters (*p*_app_ =0.9, *α · f_m_* =0.95, *η_as_* =1, trace uninfected = true). Top: The fraction of infectious (green), quarantined (blue) and cumulative exposed (yellow) people for each day of the simulated outbreak. Curves for only 20 out of the 50 runs are shown to improve legibility. Outcomes from one selected run are drawn as bold lines. Middle: *R_e_* each day is shown for the MC run drawn in bold in the top plot. The red vertical line indicates the time when 0.4% of the population have been infected, which is when interventions (other than quarantining of reported symptomatic cases, which is enabled from the beginning) are turned on. To measure their effectiveness, *R_e_* is averaged over 18 days (red area), starting 10 days after interventions commence. Bottom: Outcomes from the 50 MC runs, such as the maximum fraction of the population quarantined, are histogrammed to show the statistical variation more clearly.

The *R_e_* shown for each day is given as the average number of people infected by everyone who recovered on that day. After the interventions are turned on, *R_e_* begins to decrease and in the absence of non-linear effects reaches a plateau after approximately 10 to 14 days. In runs where more than a few percent of the population has been exposed at that time, *R_e_* declines naturally due to an increasing chance that contact persons are already infected or recovered, and therefore cannot be infected again. When reporting the *R_e_* for a simulation run, *R_e_*(*t)* is averaged in the time span of 10days to 28days after interventions start, or from 10 days to the day more than 50% of the population has been exposed, whichever period is shorter. This time window is a compromise between being far enough away from the start of interventions for the effect of the interventions to fully manifest, and not getting to close to the region where *R_e_* changes naturally. The *R_e_* reported for a scenario is the average *R_e_* over all the simulation runs for that scenario.

We consider the following outcomes:

- The fraction of the population exposed after one year of continuous interventions. The one year is counted from the day interventions start.
- The fraction of the population sick on the day when most people are sick.
- The average fraction of the population in quarantine each day over one year of continuous interventions.
- The fraction of the population in quarantine on the day when most people are in quarantine.
- The effective reproductive number after interventions.
- The fraction of simulations that did not generate an outbreak, where an outbreak is defined as at least 0.04% of the population becoming exposed in runs where interventions do not start on day 0, and is defined as at least 50 people becoming exposed in runs where interventions start on day 0.

## 4 Results

We highlight three outcomes for select scenarios and as function of the app coverage. Unless stated otherwise, the values printed in black in Tab. 1 are used for those parameters not explicitly varied in the figures or stated in the figure captions. The full set of outcomes for all scenarios is shown in the supplementary materials.

### 4.1 The effect of instantaneous contact tracing on an ongoing epidemic

Fig. 7 and Fig. 8 show three outcomes each for the four simulated symptom/reporting fractions and for *R*_0_ =3 (Fig. 7) and *R*_0_ =2 (Fig. 8). Results are in each case shown for the realistic case where tracing identifies contacts regardless of their infection status, and for the case where only infected contacts are traced. The latter is included so that results can be compared to other models, and because the difference in the number of quarantined people between the two cases indicates how many healthy people are quarantined when uninfected contacts are also traced. *R*_0_ = 2 is likely too optimistic, however the results are also valid in the situation where R was lowered to R=2 by other interventions, such as mask wearing, before tracing and quarantining starts.

**Figure 7:**
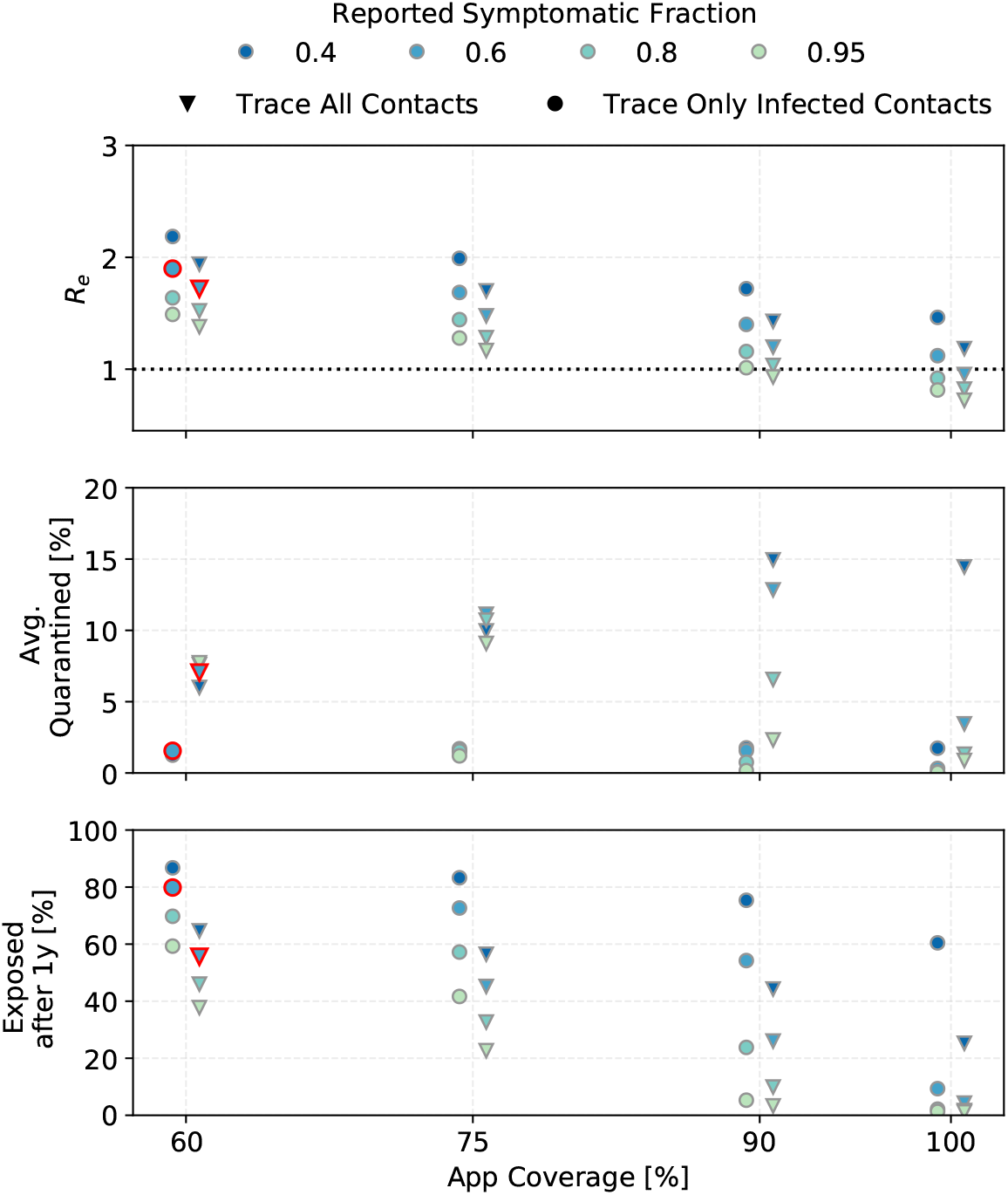
The effect of *α · f_m_* (tested symptomatic fraction) for different app coverages on the reproductive number after interventions (DCTS and quarantine) (top), the average of daily quarantined people as a percentage of the total population (middle) and the percentage of the population exposed after 1 year of continuous interventions (bottom). The points for 60% app coverage and 60% symptomatic fraction, outlined in red, will be studied further. This is for *R*_0_ =3 and *η_DCT_* = 1.

The *R_e_* shown in the top panels of Fig. 7 and Fig. 8 should be compared to Fig. 3. For example for *R*_0_ = 3 and *α* · *f_m_* = 0.6, just quarantining reported symptomatic cases yields R*_e;q_* = 2.2, so DCT only lowers *R_e_* by another 0.5 (if tracing is independent of infection status), or 0.3 (if tracing finds only infected contacts).

**Figure 8:**
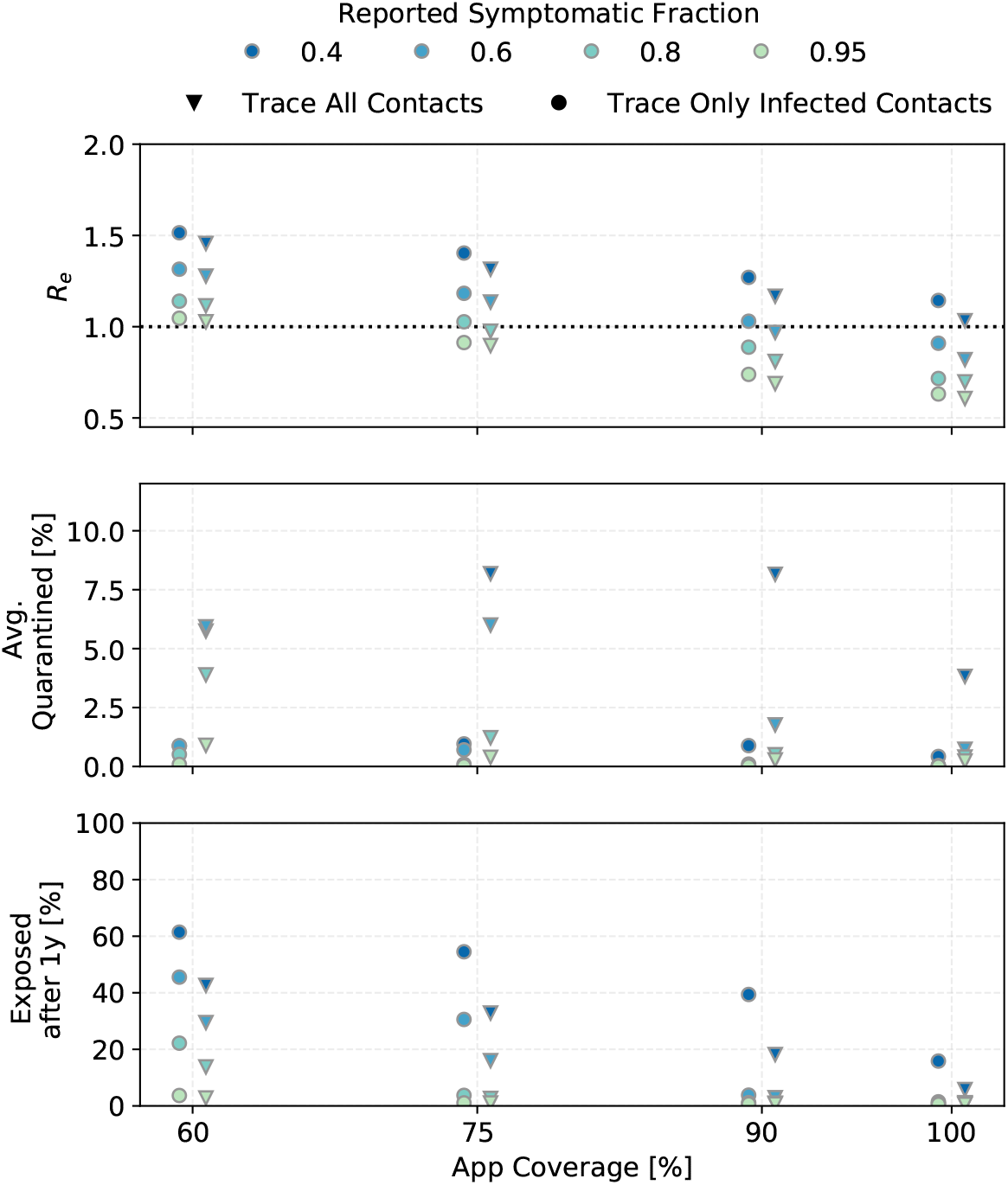
The same as Fig. 7 but for starting conditiosn where R=2. R=2 could be achieved by interventions other than DCT.

Assuming that a is about 60% in European populations and that not everyone who has symptoms sees a doctor or is tested, the region between *α* · *f_m_* = 0.4 to 0.6 is likely relevant for Europe. If the reports of higher *α* in Asian countries are due to true differences in symptom fraction rather than to differences in study methods, the higher *α* · *f_m_* values simulated should be more relevant to Asian countries.

We will use *R*_0_ = 3, *p*_app_ = 0.6, *α* · *f_m_* = 0.6, and *η_DCT_* = 1 (points outlined in red in Fig. 7) as defaults.

In Fig. 9, the lightest points correspond to these defaults. The other colors indicate what happens when the tracing efficiency is reduced. For the lower app coverages, the results are barely sensitive to *η_DCT_* because DCT is not very effective to begin with.

**Figure 9:**
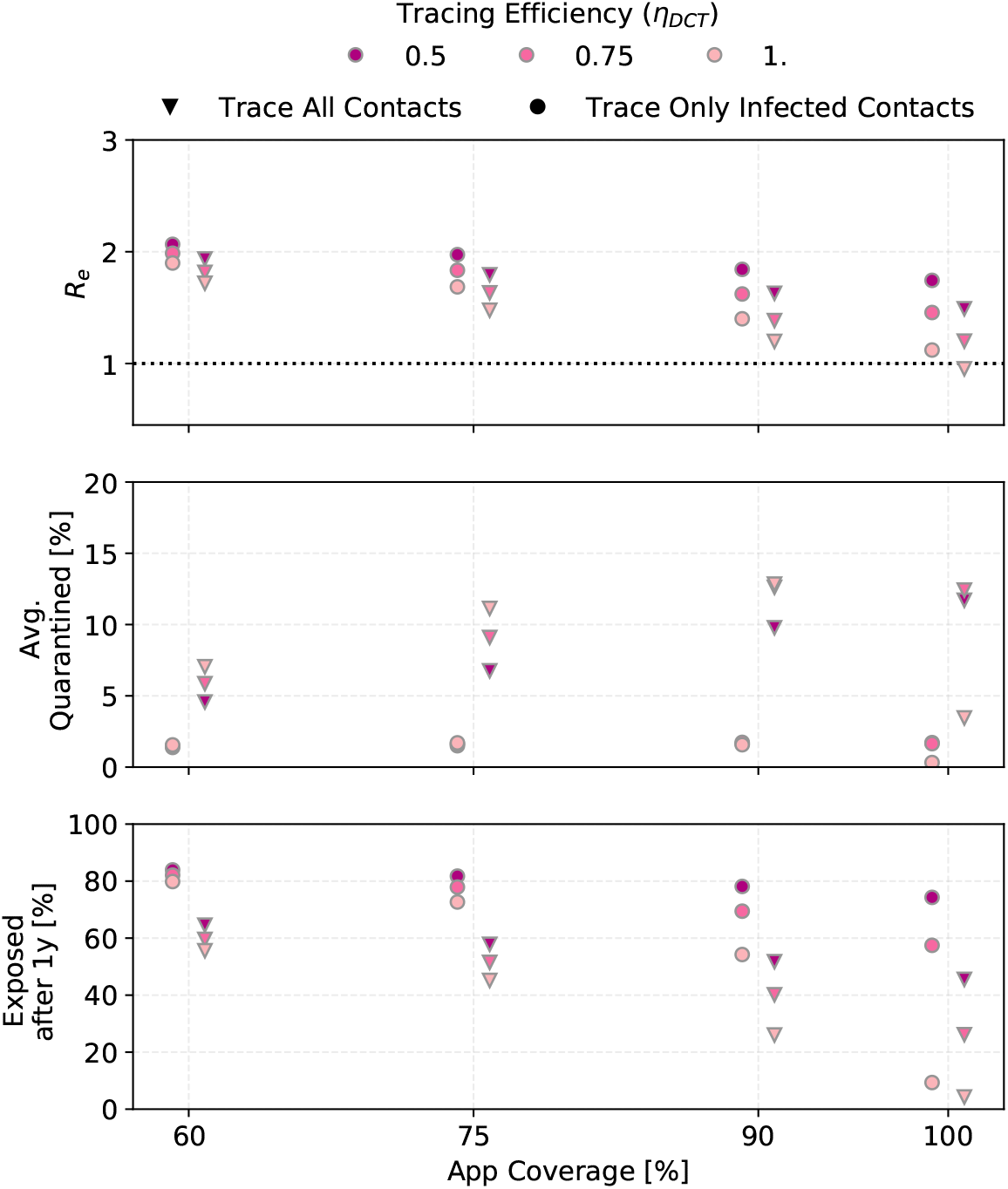
The effect of the tracing probability *η_DCT_* for different app coverages (*R*_0_ =3, *α* · *f_m_* =0.6).

In the realistic case where traced uninfected contacts are quarantined until two test results are negative (see Sect.2.3), as many as 15% of the population are in quarantine on each day of the simulated outbreak, most of them healthy. Without a POC test to release healthy contacts, this number rises to 25%. At the peak of the outbreak, approximately 30% of the population is quarantined and half of those quarantined are actually sick.

People are not available to be infected while in quarantine, so the mean number of contacts per day, and with it the effective reproductive number, goes down and fewer people become exposed. The number of people quarantined rises with higher app coverage (because more people are traced in that case) and with a higher number of people exposed (because there are more index cases). A higher app coverage eventually leads to fewer exposed people though. Hence for a given reported symptomatic fraction, the number of people quarantined rises until an app coverage of approximately 75% (for lower reported symptomatic fractions) or 90% (for higher reported symptomatic fractions) and then falls sharply.

Contact tracing cannot reduce R below 1 in any of the simulations presented here except for *α · f_m_* ≥ 0.8 and *p*_app_ ≥ 0.9 (if *R*_0_ = 3) or *p*_app_ ≥ 0.7 (if R when tracing and quarantining is started is 2), and perfect tracing probability.

We note that in some cases, the fraction of the population exposed after 1 year is higher than the herd immunity level. The herd immunity level is defined as the fraction of the population that must be immune for the increase in new infections to not be able to grow exponentially, that is for *R_e_* to become 1. In an ongoing epidemic, many people are infectious when this point is reached, and the number of exposed people continues to rise until enough people are immune for *R_e_* = 0, therefore the curve overshoots herd immunity level.

### 4.2 Contact tracing in combination with random testing and social distancing

To control the epidemic, R must be reduced by additional measures. We simulated the effect of random testing (RT) and social distancing (SD). Fig. 10 shows the outcomes for our standard scenario with the addition of RT of 5% and 20% of the population per day, and social distancing bringing *n_c_* to 0.8 and 0.6 of its original value. The reduction in contact rate is always connected to an upper limit in the number of contacts as shown in Table. 1.

**Figure 10:**
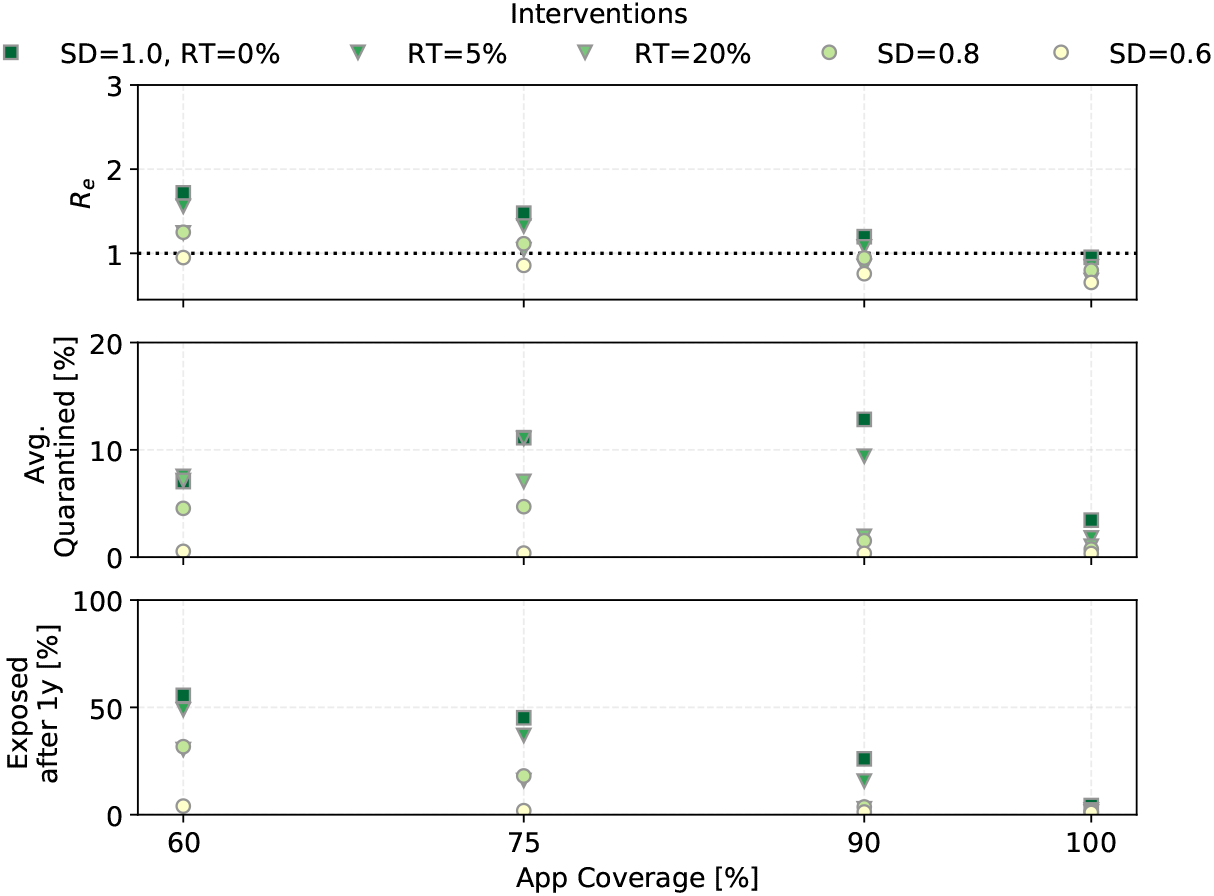
The effect of social distancing (SD) and random testing (RT) for different app coverages in combination with CT (*R*_0_ =3, *η_dct_* =1, *α · f_m_* =0.6).

Random testing even at 20% of the population per day in combination with contact tracing can only achieve *R_e_* ≤ 1 for *p*_app_ ≥ 0.75. It does however bring *R_e_* close enough to 1 to significantly reduce the fraction of the population that becomes exposed, even for lower app coverages.

Social distancing reliably reduces the reproductive number. Social distancing to just 80% of the contact rate does as well as randomly testing 20% of the population each day. Reducing the contact rate to 60% pushes *R_e_* below 1 for 60% app coverage.

### 4.3 The effect of reduced contagiousness of asymptomatic carriers

As we saw in Fig. 3, in a situation where reported symptomatic cases are quarantined, down-scaling the contagiousness of asymptomatic carriers reduces R significantly. Fig. 11 shows the outcomes when DCT is then applied.

**Figure 11:**
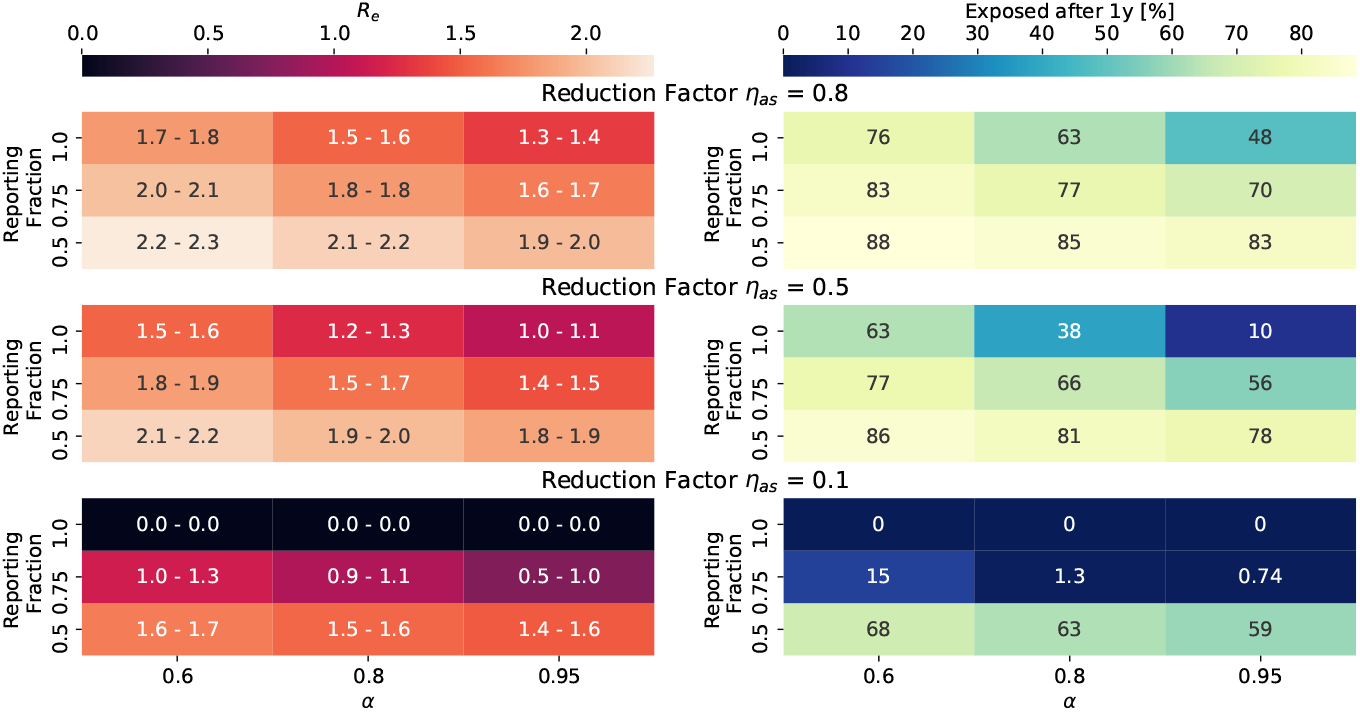
Outcomes when the contagiousness of a- and pre-symptomatic carrieres, *η_as_*, is smaller than 1. Settings are *R*_0_ =3 *p*_app_ =0.6, *η*_DCT_ =1.0, trace uninfected contacts = false. The values printed for *R_e_* correspond to: (first number) the mean minus the standard deviation, and (second number) the mean plus the standard deviation, of the distribution of *R_e_* from 100 simulations (compare Fig. 6 bottom right panel), while the color of the field shows the mean. Where values are exactly 0.0, none of the 100 simulations had an outbreak (compare Sect. 4.5).

In the case where *η_as_* =0.1 and all symptomatic carriers are reported when symptoms start, the simulations generated no outbreaks (fewer than 400 people became infected in total). When 75% of symptomatic cases are reported, *R_e_* has large fluctuations between simulation runs, and outcomes are very sensitive to the fraction of symptomatic cases.

### 4.4 The effect of timing, delays, and second order tracing

Fig. 12 shows the outcomes as a function of tracing delay, that is the time in days between when an index case is identified and when his or her contacts are traced and quarantined. Outcomes are again grouped by whether or not uninfected contacts are identified by tracing. Results are also shown for both 1st order tracing and 2nd order tracing and for the two incubation time curves (the default one with mean incubation time (IT) of 7 days and the alternative one with the shorter IT=4 days). The yellow marker uses the default incubation time curve with the alternate transmission probability curve (IC) where there is less pre-symptomatic transmission. In addition to the simulation results, the calculated *R_e_* from the age-of-infection model is shown for the settings using the default incubation time and transmission probability curves, and first order tracing. Approximations had to be made in the calculation, hence the absolute value is not expected to match the simulation results perfectly.

**Figure 12:**
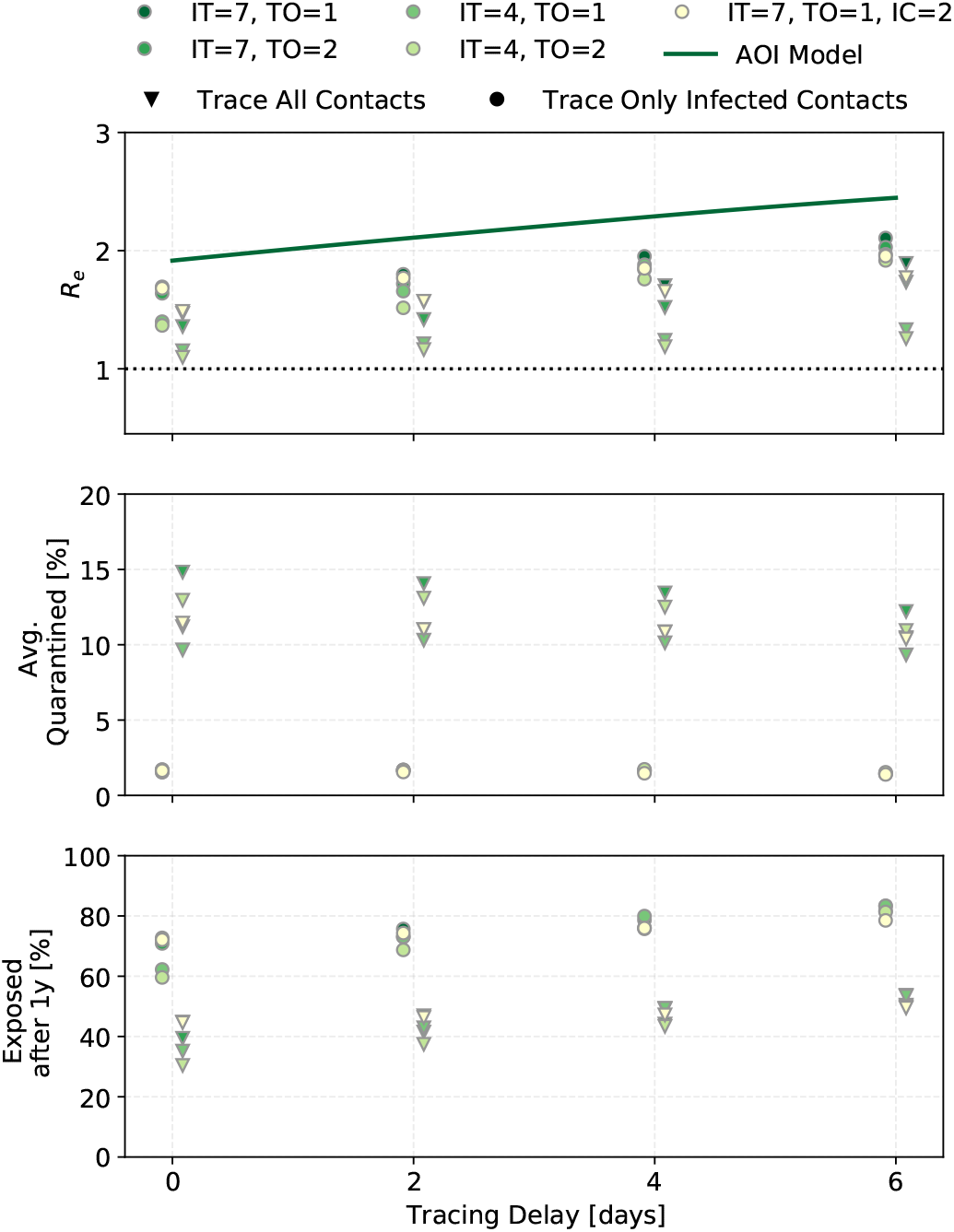
The effect of incubation time (IT) and tracing order (TO) for different tracing delays. IT=7 refers to the curve with mean incubation period of 7 days, and IT=4 to the one with mean incubation period of 3.6 days. The yellow point shows results for the alternate transmission probability curve (IC) as shown in Tab. 1 (grey values). Predictions from the “age-of-infection” (AOI) model, where only infected contacts are traced, are shown as the dark green line for parameters IT=7, TO=1 - the result shown is not exact (see appendix). This is for *p*_app_ = 0.75, *η_DCT_* = 1, and *α · f_m_* = 0.6.

Results are shown for an app coverage of 75%. Some of the dynamics are quite sensitive to the app coverage (see appendix), and at 75% trends are clearer than at 60% app coverage.

The difference between first and second order tracing is small in all three outcomes (this changes in some situations for higher app coverages). For the default incubation time curve with mean of 7 days, tracing delays of up to 6 days have only a small effect, increasing the number of people exposed after 1 year from approximately 72% to 82%. When the mean incubation time is only 4 days, tracing delays more strongly reduce *R_e_* The infection probability curve with less pre-symptomatic transmission probability only very slightly improves the outcomes, though the effect becomes bigger with increasing tracing delays.

Second order tracing can find the infector and through him or her, the ’siblings’ of the index case. The chance that the infection took place within Δ*T_trace_* is higher with a longer Δ*T_trace_*. However even for Δ*T_trace_* =14 days, the outcomes are not significantly different. The look-back time must be balanced against the number of healthy people quarantined. People typically become index cases before they have recovered, and thus would have had a chance to infect others in the approximately 7 days prior. Looking back longer than that means one has a bigger chance of finding the infector, but it also means tracing many uninfected contacts.

When considering the realistic case where uninfected contacts are traced, second order tracing with a 7 day look-back time sends about 1.3 times as many people into quarantine on average over 1 year as first order tracing.

### 4.5 Outbreak probability

The results discussed so far consider situations where an outbreak is ongoing and interventions are started at some point into the outbreak. But not all simulation runs result in an outbreak. The stochastic nature of the outbreak means that there are large statistical variations at the beginning of the chain. For example, if patient zero happens to not infect anyone, no outbreak happens.

The chance for an outbreak to occur increases with R. The more people a case typically infects, the less likely it is that cases at the beginning of the infection chain do not infect anyone. Therefore, keeping interventions in place even in populations without an ongoing outbreak can be useful to decrease the probability that an outbreak will occur when a case is introduced into the population, for example through travel.

Figure 13 shows the outbreak probability as a function of the reproductive number when an infected person enters a fully susceptible population.

**Figure 13:**
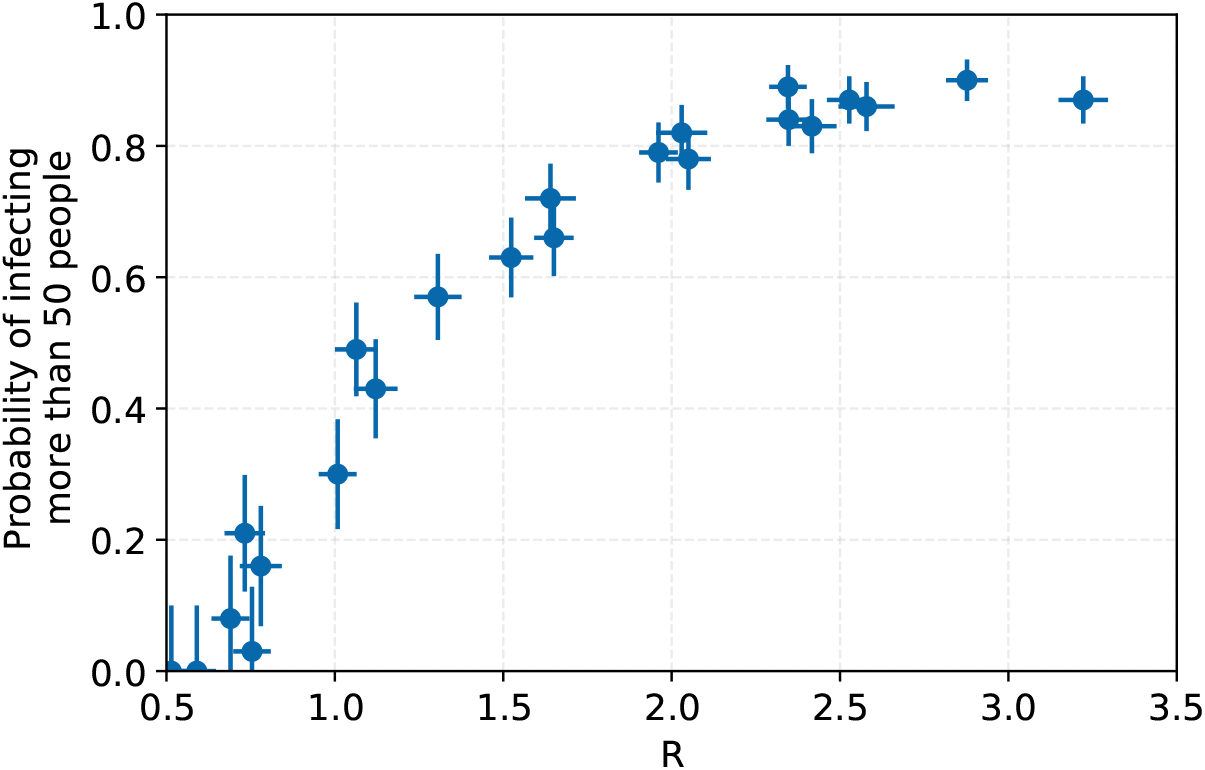
The probability for an outbreak to start as a function of the reproductive number at the time when patient 0 enters a fully susceptible population. An outbreak here is defined as more than 50 people becoming infected. The error bars shown are statistical.

### 4.6 Sensitivity of results to the social contact structure

The results presented so far assumed a homogeneous population of size 1 × 10^5^ and a distribution of the number of contacts with infection potential per day from Eq. (3). We also ran some sets of parameters for different population sizes and for different contact structures. The results are shown in Fig. 14.

**Figure 14:**
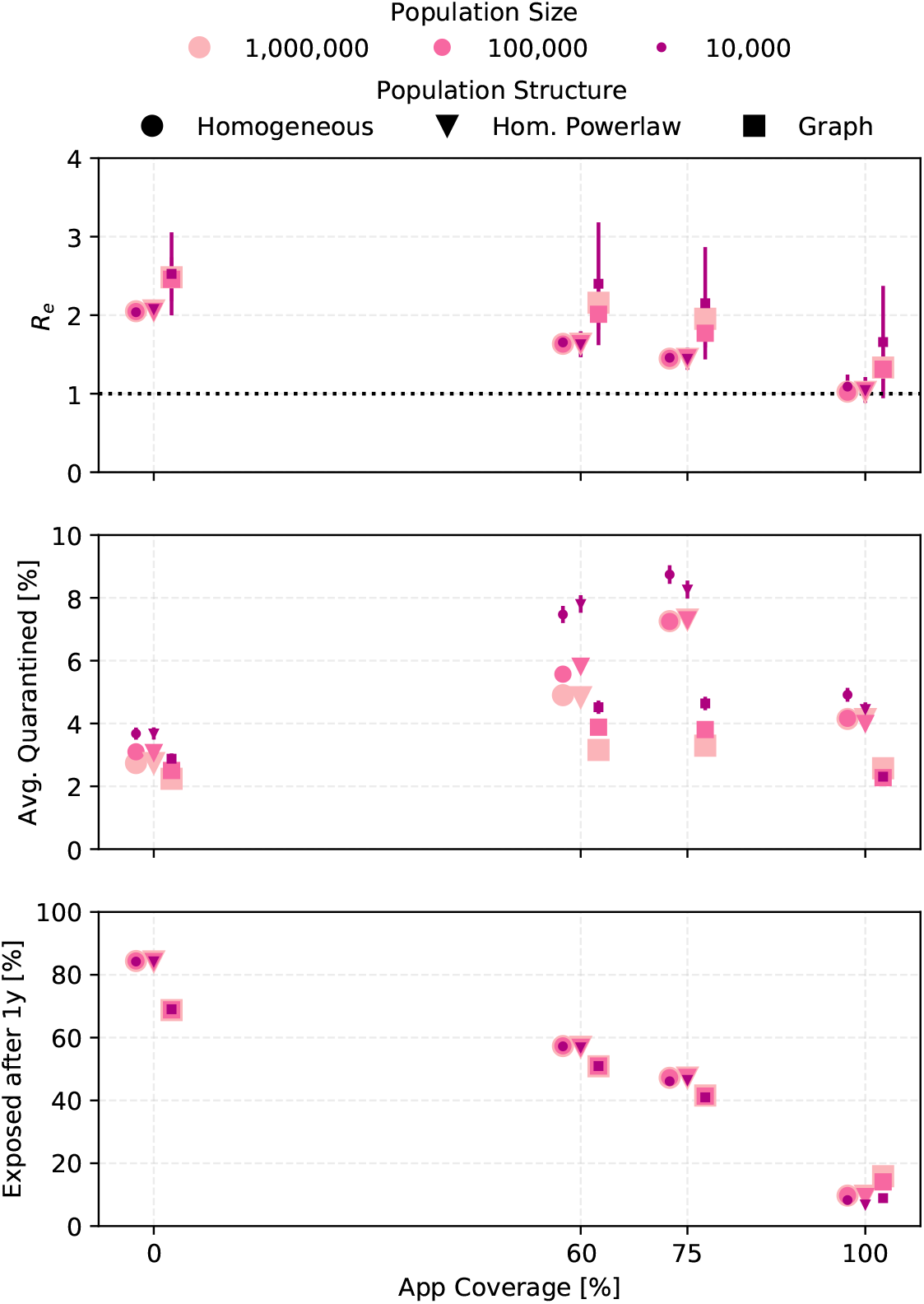
The sensitivity of the outcomes to the size and the structure of the simulated population is shown. The three population structures (described in Sect. 2.1) correspond to the homogeneous population with the gamma distribution (Eq. (3)) describing the number of contacts per day, a homogeneous distribution using the power law (Eq. (4)) for the number of contact per day, and the social graph population. Settings are *R*_0_ = 3, *α · f_m_* = 0.6, *η_dct_* = 1, trace uninfected=true. For the 10 000 people population, outcomes have large statistical fluctuations and error bars show the error on the mean. For the other points, the error bars are smaller than the marker size.

The introduction of a social graph introduces non-linear effects that change *R_e_* on timescales much longer than what is captured by our standard analysis. In some cases, this means that fewer people are exposed after one year, even though *R_e_* is higher (see Fig. 21 in Appendix C).

## 5 Discussion

Contact tracing relies on index cases from which to trace. When there is a large fraction of mildly symptomatic and asymptomatic carriers who never go to the doctor or get tested, many carriers do not become index cases, so DCT does not have a large impact. Outcomes improve strongly the higher the fraction of reported symptomatic carriers. This is partially because DCT is more efficient, and partially because R is additionally reduced just from quarantining the index cases. Therefore it is crucial that every person with even the mildest symptoms has easy access to a COVID-19-test.

The extend to which pre- and asymptomatic carriers drive the outbreak depends on their contagiousness. If for some reason they are less contagious than symptomatic carriers, missing them as index cases does not worsen outcomes much. In the case where *η_as_* is 0.1, as proposed for example in [4], quarantining of index cases, without CT, reduces R from *R*_0_ = 3 to *R_e_* < 1 even when just 40% of cases are symptomatic.

Randomly testing a fraction of the population regularly to find unreported carriers helps to make up for the large fraction of asymptomatic carriers. We find that a very large fraction of the population must be tested daily to significantly improve outcomes. For our default parameters, even when testing 20% of the population daily, at least 90% of the population would have to use the DCTS for *R_e_* to become smaller than one.

Reducing the contact rate (social distancing) by as little as 20% is as effective as testing 20% of the population every day while requiring fewer people to be quarantined.

Tracing delays of a few days do not significantly worsen the outcomes. Two studies, Ferretti et al. [4] and [5], indicate that a DCTS could control a SARS-CoV-2 outbreak (that is achieve *R_e_* < 1) because it allows for contact tracing without delays. We find that the asymptomatic infectiousness scaling of *η_as_* =0.1 used by [4] is the main driver of their *R_e_* and given these starting conditions, DCTS only has to lower R by a small amount to achieve outbreak control and is therefore then effective. Kretzschmar et al. [5] are more careful about the reduction in R achievable with DCTS, but do confirm the improved outcomes with short tracing delays. However, [5] use a very short latency period. With the longer median latency periods consistent with recent large-scale studies, this effect is small. Therefore, the advantage of a DCT in the case of COVID-19 lies mostly in the possibility to scale tracing to a large number of cases without needing a large increase in the number of manual contact tracers.

Most models consider that contacts that were actually infected are traced with some probability. In reality, it is impossible to tell immediately whether or not a traced contact has been infected. Even if a test performed immediately on tracing is negative, it could just mean that the person is still in the latent period. Therefore, all traced contacts should be quarantined and tested multiple times. In principle, one could devise other schemes, such as testing each traced person every morning (e.g. with a POC tests that can be done at home and gives results within minutes) for a few days without requiring quarantine unless the test comes up positive. Right now, such frequent testing is not realistic in most countries.

We find that including the effect that quarantining of uninfected contacts has on the outbreak dynamics can lead to significantly different, typically more positive, outcomes compared to models where this effect is ignored. The improvement in outcomes is due to the large number of people quarantined even though they are healthy. Our simulations probably underestimate this number, because we use contact rates for the types of contacts that have a high chance of transmitting a respiratory virus. A DCT system will typically pick up many contacts who were in spacial proximity to the index case, but not in a manner that was likely to transmit the virus, so the number of contacts traced per index case could be bigger in reality. For any serious large-scale use of a DCT system during an ongoing epidemic, dealing with these uninfected contacts in quarantine is going to be a major challenge, especially as the compliance of the population with quarantining procedures may decrease once someone has been traced and quarantined multiple times.

The statistical nature of virus transmission and contact rates leads to large variations in outbreak dynamics at the start of the outbreak. Sometimes, an infectious person entering a susceptible population does not start an outbreak. This becomes less likely the higher R is. This also means that under identical conditions, one population could have hundreds of cases within a week of the arrival of patient 0, while in another population the case number does not start rising for several weeks, just by luck.

Beside control of the outbreak, that is achieving *R_e_* below 1, an important outcome is how many people will have been exposed by the time a vaccine might be available. Due to the heavy social and economical burden imposed by virus control, some countries are aiming at *R_e_* around 1, rather than total control. Our simulations assume that the same interventions are applied throughout the epidemic, so the outcomes over 1 year are guidelines rather than realistic predictions for any real country. They do give a qualitative idea of what achieving a given *R_e_* means in terms of the number of people exposed (and with that, the number of fatalities).

For reasons of computing power, most simulations were run for a homogeneous population. In reality, society is organized into social units. Introducing such social units into the simulation means that an infectious person tends to meet the same people every day, exposing them again and again. This leads to slight changes in outcome, while the qualitative results remain the same.

### 5.1 Limitations

We assumed that all people, once they have recovered from the infection, are immune to a secondary infection. Whether and for how long a recovered person is immune remains to be answered. Studies show that neutralizing antibodies are produced during infection and to a higher degree in symptomatic carriers, but decline significantly 2-3 months after recovery [54, 55]. The minimal antibody titer to confer protection is, however, still unclear. Furthermore, memory T cells to SARS-CoV-2 have been found in patients including asymptomatic and mildly symptomatic ones, which likely contribute to protective immunity as well [56].

In our models, everyone adheres to quarantine protocols. That is, every time someone is alerted by the DCT system to having been in proximity of a contagious individual, this person must follow the quarantining and testing procedure. This is crucial to suppress pre- and a-symptomatic transmission, but may be difficult to achieve in reality. We also assume that the fraction of symptomatic individuals who see a doctor/get tested do so the day they become symptomatic.

The transmission probability in our models changes with the time since infection, but not between individuals. Current research, however, suggests that COVID-19 is overdispered, meaning some individuals spread the virus to many others, in so-called “superspreading events”, while most do not transmit the virus at all or only to very few people [57]. Part of this overdispersion is due to the random nature of the contact number - some people just meet more others, and is therefore included in our models (see Sect. 2.2 and Fig. 2).

We assume that no manual tracing is performed at all. Typically, the types of close contact persons to whom spreading the disease is most likely, that is friends and family, can be manually traced without much effort, hence the fraction of infected contacts traced could be larger in reality.

## 6 Summary

Many countries enforced a policy of ’shelter–in–place’ and/or extreme social distancing, effectively putting most of the population into quarantine. This significantly slowed down the infection rate [58][59], but came with large economical and social costs to society. World-wide, a lot of effort has been put into the development of CT systems, in the hope that large-scale CT could replace other public health measures at much smaller cost to society.

We modelled the effect of instantaneous DCT in combination with a testing and quarantining protocol, as well as random testing and social distancing, on an ongoing COVID-19 epidemic. Results were validated by running the scenarios with two independently developed individual-based models, which were further cross-checked by two types of deterministic models. We modelled many different parameter values for the still not well-known properties of SARS-CoV-2, COVID-19 and for the interventions, leading to well over 10 000 simulated scenarios. The goal was to find the regions in this parameter space where CT without additional interventions could lower the effective reproductive number enough to halt exponential growth.

Wherever modelling approximations had to be made, we chose defaults that lead to better outcomes, hence these results are likely on the optimistic side. Our results are stable under different simulated social structures and epidemiological parameters, with significantly different outcomes seen only when varying the fraction of asymptomatic individuals or down-scaling the contagiousness of pre- and asymptomatic cases.

We find that for large regions of the parameter space, including the currently most likely parameter values, an outbreak of COVID-19 cannot be fully controlled by DCT even if a large fraction of the population uses the system. Furthermore, if interventions are started once an outbreak is already ongoing, DCT causes a large fraction of the healthy population to be traced and quarantined.

DCT can be combined with other measures, such as face-mouth coverings, social distancing, and/or random testing, to achieve outbreak control.

The availability of fast testing, and coordination of test results with the DCT system, are crucial to allow symptomatic cases to become index cases for tracing, and to release traced healthy contacts from quarantine. Since SARS-CoV-2 symptoms are unspecific, everyone with even a slight cough of fever must be able to get a test (a) quickly, because the infection probability peaks just before symptom onset and then falls quickly and people who are not sure they are infected likely will not effectively quarantine themselves, and (b) easily, so that a large fraction of symptomatic cases do seek out testing. The gains of a DCT system in outbreak control quickly vanish if many symptomatic cases do not seek out testing, or if positively tested individuals do not become index cases.

## Data Availability

The code used to produce the simulations will be available on GitHub.

## 7 Acknowledgements

We acknowledge the support by the DFG Cluster of Excellence ’’Origin and Structure of the Universe”. The simulations were carried out on the computing facilities of the Computational Center for Particle and Astrophysics (C2PAP) and on the Max Planck Institute for Physics computing cluster. This work was undertaken in the context of the ContacTUM collaboration.

## A The ODE model

The compartment model is based on the SEIR-type model from Reference [53] and is extended to visualize infectious individuals for their whole infectious period *T*_con_ even though they might be quarantined or hospitalized (see Figure 15). To do so, we introduce the convalescing compartment ’*C*’ in between the infectious *I* and recovered *R* status.

**Figure 15:**
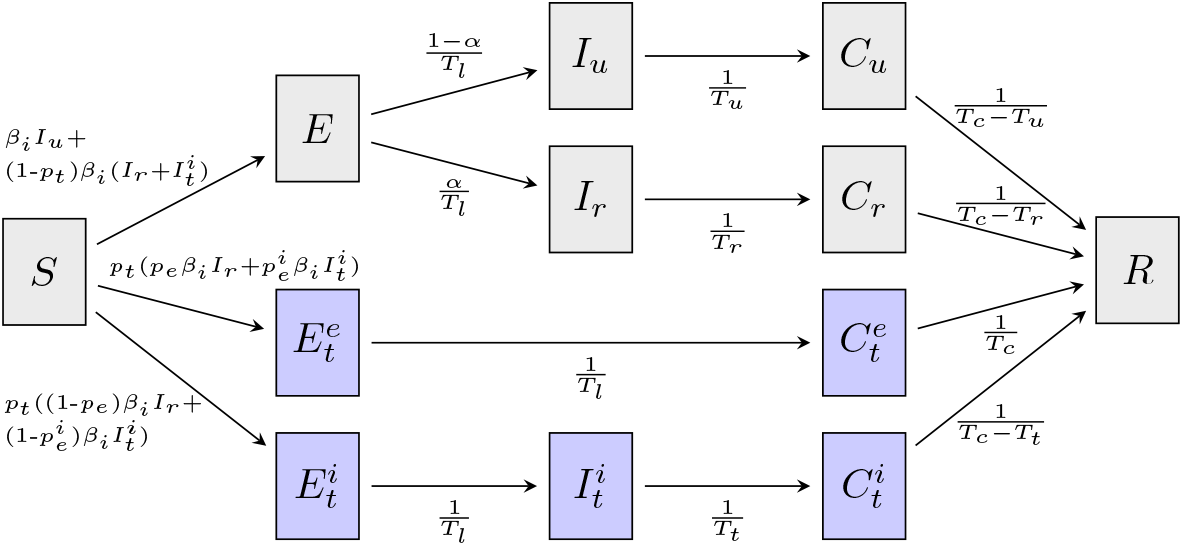
Structure of the compartmental model. The model is based on a SEIR-type model from Ref. [53] and is extended for the convalescing (*C*) compartment. A person in the convalescing status is technically still infectious, but the chance to infect further is drastically reduced, as this person is either isolated or the viral load is effectively too low. The sum of the specific infectious periods is given by *T*_Con_. Untraced compartments are indicated in gray and traced compartments in blue. See Table A for the definitions of the variables.

*I* denotes the infectious individuals that are able to infect susceptible individuals *S* for the mean infectious time *T*. In contrast, *C* are convalescing individuals that are still infectious but cannot infect further as they are either isolated or the probability to infect is effectively too small. The latter case reflects the fact that the probability to infect is reduced for large times post infection as shown in Figure 1.^1^ After the residual infectious time *T*_con_ — *T*, *C* enter the recovered status R. In Fig. 18, the infectious population is defined as the sum of all *I* and *C* subgroups.

**Figure 16:**
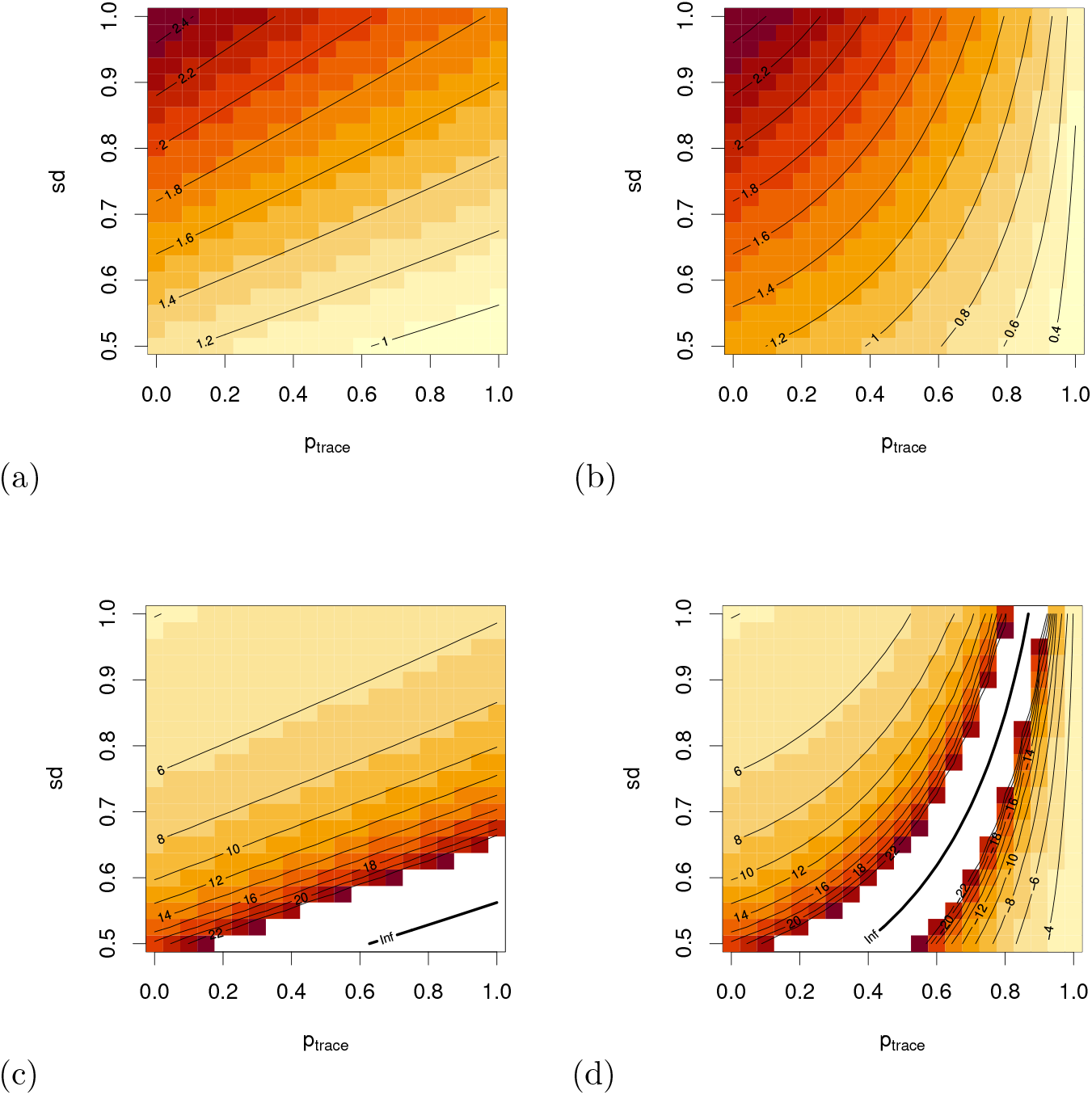
Influence of social distancing (reduction of the contact rate) and contact tracing on *R_e_* and *T*_2_. (a) and (c): One-step tracing, (b) and (d) recursive tracing. (a) and (b) *R_e_*, (b) and (d) *T*_2_. Note that in (b) and (c), there is a singularity for *T*_2_ at the line *R_e_* = 1, where *T*_2_ becomes infinite.

**Figure 17:**
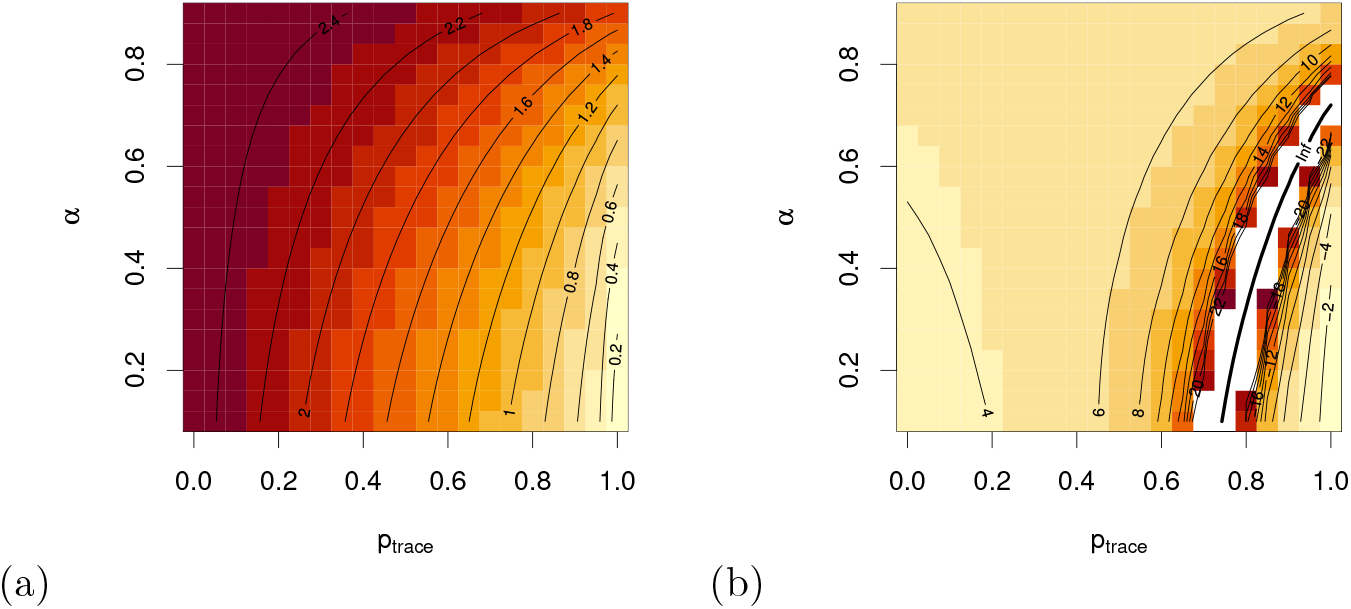
Influence of the assumed fraction of asymptomatic and unreported cases on (a) *R_e_* and (b) *T*_2_, where we also take *η_DCT_* into account (only recursive tracing considered).

**Figure 18:**
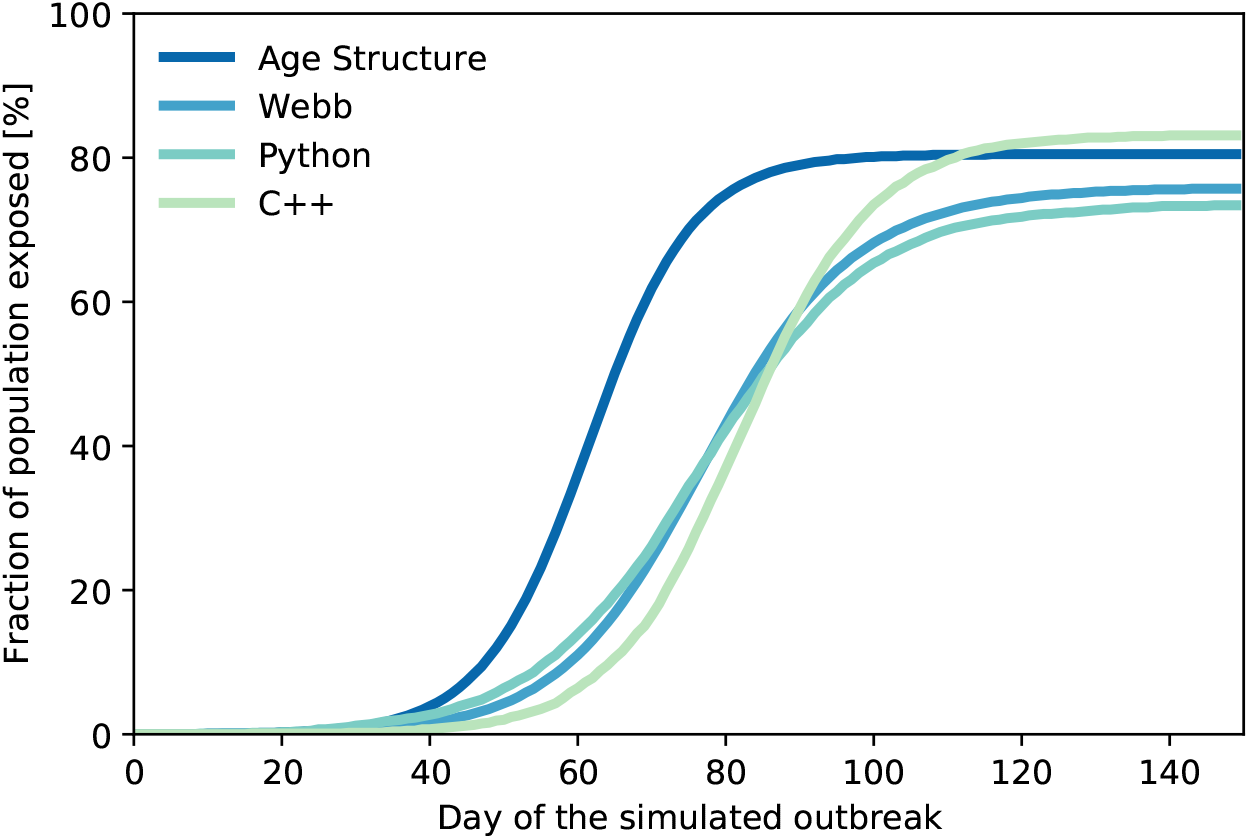
The cumulative number of exposed people from the four models and for the settings from Tab. 3.

The model takes into account contact tracing and differentiates between untraced and traced individuals. Contact tracing is triggered by reported infectious individuals that appear with the fraction *α* of the untraced individuals. The model incorporates forward contact tracing, e.g. one traces the contacts that have been infected by the reported index case and predicts the probability *p_e_* that the traced contact is still exposing when traced. This prediction relies on exponential probability distributions for the latent and infectious period. We assume that traced contacts that are still exposing 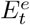 are immediately isolated. In comparison, contacts that have been traced during their infectious period 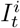 could have already infected further people. The corresponding mean infectious time *T_t_* of 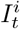 is calculated within the model and depends on the latent and the infectious period of the reported infectious individuals. Traced infectious contacts 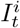 can trigger contact tracing as well and the probability that the traced contact of 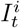 is still exposing is given by 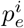 that depends then on *T_t_*. Table A summarizes the parameters and definitions of the compartmental model. For a detailed description and the proof of the computation of *p_e_*, 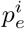, and *T_t_*, we refer the reader to Reference [53].

In order to compute the doubling time *T*_2_ and the basic reproduction number *R*_e_, we focus on the initial infection-free state, in which the entire sample population is given by *S*_0_ [53, 60]. At this stage, the system of differential equations can be linearised, and the aforementioned parameters can be extracted from the Jacobian of the system *J*. In particular, the doubling time is computed as ln(2)/λ, where λ is the largest eigenvalue of *J*, known as the exponential-growth parameter. On the other hand, the basic reproduction number is obtained using the *next-generation analysis*, in which the Jacobian is split into two matrices: *F*, containing the terms relative to the generation of new infections, and *V* containing the transfer from one infectious compartment to another. The basic reproduction number *R_e_* is then obtained as the spectral radius of the matrix *FV*^−1^.

We derive the mean latent and infectious times *T*_lat_, *T*_r_, and *T*_u_ using the probability functions described in Section 2 to 5.0 d, 1.4 d, and 2.5 d, respectively. Further, the total infectious time T_con_ is assumed to be 10.4 d.

The model is given by the following set of ordinary differential equations

(ODEs):

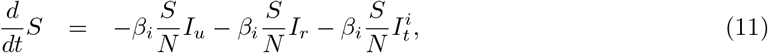

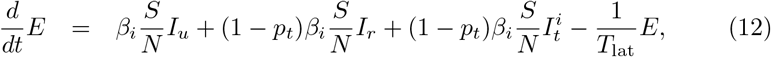

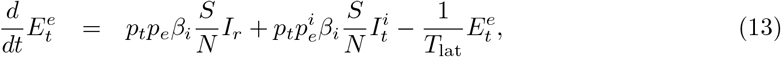

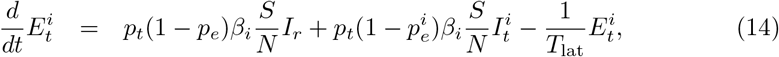

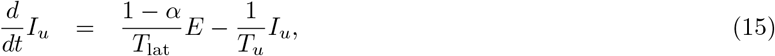

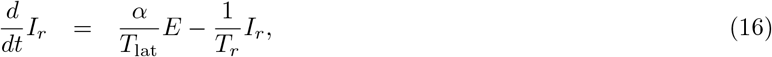

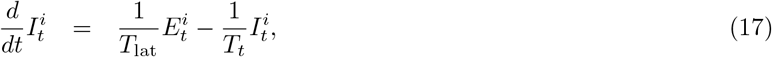

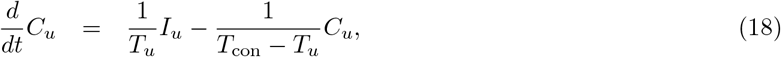

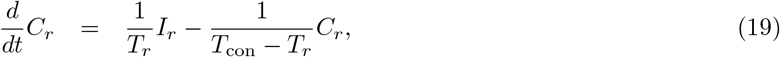

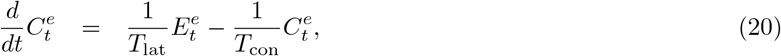

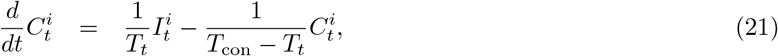

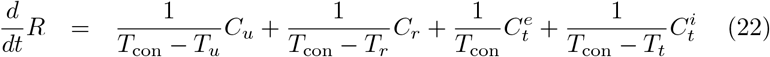

**Table 2:** Parameters and Definitions of the compartmental model.

| Parameter | Definition |
| --- | --- |
| $S(t)$ | number of susceptible individuals |
| $E(t)$ | number of untraced exposing individuals |
| $E_t^e(t)$ | number of exposed individuals that will be traced during exposed phase |
| $E_t^i(t)$ | number of exposed individuals that will be traced during infectious phase |
| $I_u(t)$ | number of unreported infectious individuals |
| $I_r(t)$ | number of reported infectious individuals |
| $I_t^i(t)$ | number of infectious individuals that will be traced during infectious phase |
| $C_u(t)$ | number of unreported convalescing individuals |
| $C_r(t)$ | number of isolated convalescing reported individuals |
| $C_t^e(t)$ | number of isolated convalescing individuals that were traced during exposed phase |
| $C_t^i(t)$ | number of isolated convalescing individuals that were traced during infectious phase |
| $R(t)$ | number of recovered and immune individuals. Note that individuals in $R$ could also be dead. |
| $N(t) = N$ | constant sum of all individuals |
| $\beta_i$ | transmission rate |
| $\alpha$ | reporting fraction of all individuals. Note that we do not distinguish between asymptomatic and symptomatic cases. |
| $T_{\text{lat}}$ | latency period |
| $T_{\text{con}}$ | total infectious period |
| $T_u$ | infectious period of unreported individuals |
| $T_r$ | infectious period until reporting |
| $T_t$ | infectious period of traced individuals $I_t^i$.<br>$1/T_t = 1/\bar{T} + 1/\hat{T}_t$ with $\hat{T}_t = \frac{1}{T_r} \int_0^{T_r} [T_r + \delta - (\int_r^{T_r+\delta} \frac{x \exp(-(x-r)/T_{\text{lat}})}{\int_0^{T_r-r+\delta} \exp(-s/T_{\text{lat}}) ds} dx)] dr$ and $\bar{T} = \alpha T_r + (1 - \alpha) T_u$ [53]. |
| $p_t$ | tracing probability. Note that we do not distinguish between $p_{\text{app}}$ and $\eta_{DCT}$ |
| $p_e$ | probability that individual is traced during exposed phase. Probability depends on $T_{\text{lat}}$, $T_r$, $\delta$: $p_e = \frac{T_{\text{lat}}}{T_{\text{lat}} + T_r} e^{-\delta/T_{\text{lat}}}$ [53] |
| $p_e^i$ | probability that individual who got infected by $I_t^i$ is traced during exposed phase. Probability depends on $T_{\text{lat}}$, $T_t$, $\delta$: $p_e^i = \frac{T_{\text{lat}}}{T_{\text{lat}} + T_t} e^{-\delta/T_{\text{lat}}}$ [53] |
| $\delta$ | time between reporting of index case and tracing of contact |

## B The age since infection model

We propose here a simple deterministic model for contact tracing, where the class of infeceteds is structured by age since infection.

Let *S*(*t*) denote the density of susceptible at time *t*, *I*(*t*, *a*) the density of infecteds at time *t* with age of infection *a*, and *R*(*t*) the removed individuals at time *t* (recovered, quarantined, dead – in any case, not infectious any more). Note that the infected individuals may not be infectious, if they are still in the latent period – exposed and infectious are distinguished by the age of infection. The total amount of the infecteds at time *t* is given by 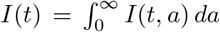. 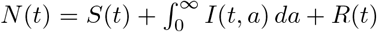 denotes the total population size. Since we do not consider population dynamics (i.e. the population size does not change), *N*(*t*) = *N* is a constant.

We first describe the model without contact tracing and discuss how to incorporate contact tracing afterwards. Infected individuals with age of infection *a* have infectivity, *β*(*a*), an infected person recovers spontaneously without diagnosis at rate *μ*(*a*); alternatively, an infected person develops symptoms and gets diagnosed at rate *σ*(*a*). In that case, he/she is quarantined immediately and will not infect further susceptibles. We chose standard incidence, s.t. the model equations become

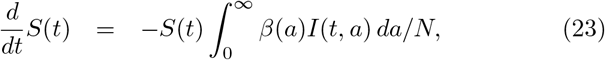

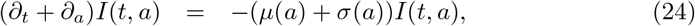

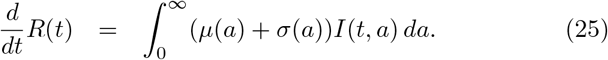

In order to prepare for the effect of contact tracing, we slightly rewrite the model equations. Thereto, we note that

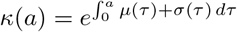

is the probability to be in the class *I* at time of infection *a*. Below, we will modify *κ*(*a*). For now, we note that

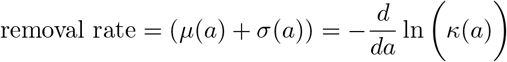

(which is also called the hazard rate). therewith, our model becomes

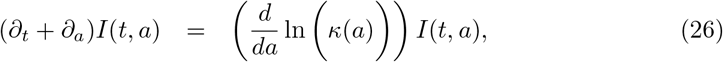

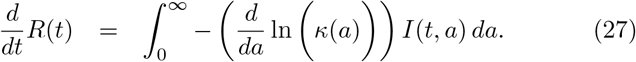

In order to compute the effect of contact tracing, we only need to adapt *κ*(*a*) accordingly. The overall model structure is not touched. The analysis of the probability to be infectious at age of infection *a* as given below is mathematically precise for the onset of the infection.

### Contact tracing

We now slightly switch the perspective, and consider single individuals. What follows was basically published in [52]; we summarize these considerations for the convenience of the reader. Below, we investigate a forest: An infected individual infects other individuals, in that, we get a random tree with a directed node from infector to infectee. Individuals who recover leave this tree, that is, we are left with a forest.

Contact tracing acts on this forest. At rate *σ* (*a*) an individual is (directly) detected/diagnosed and forms an index case. All neighbouring individuals (within the forest of infecteds) have probability p to be detected by contact tracing. Either we stop here (one step tracing) or the individuals detected by CT form recursively new index cases (recursive tracing). This process modifies *κ*(*a*): contact tracing increases the probability to be removed at age of infection a.

In order to exactly understand how *κ*(*a*) does look like, we use a mathematical fiction: First, we assume that we can only trace contacts from infectee to infector (backward tracing). Then, we assume that we can only trace contact from infector to infectee (forward tracing). Last, we combine both approaches to understand the full tracing we aim at.

### Backward tracing

#### Proposition B.0.1

*The probability to be infectious for an individual at age of infection a for recursive contact tracing follows the following system of integro-differential equations*,

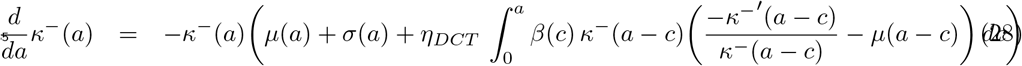

*with κ*^−^(0) = 1.

##### Proof

Clearly, without contact tracing, we have 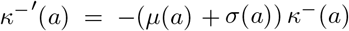. If backward tracing is active, we have an additional component of the removal rate that is caused by a tracing event triggered by an infectee. As only infectees cause tracing (tracing events are only triggered by “children”), the probability to be infectious at a given age of infection a is the same for infector and infectee. Hence the recovery rate of an infectee can be written as the hazard rate

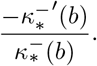

This hazard rate includes the rate of direct observation (that triggers a backward tracing event), the rate at which the infectee is discovered by recursive tracing (which triggers tracing event), and by spontaneous removal (which does not trigger a tracing event). Spontaneous removal does not lead to contact tracing, hence we subtract this rate and find the rate at which direct diagnosis or detection by contact tracing happens,

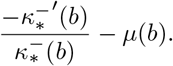

The focal individual (for which we compute *κ*^−^(*a*)) produces during his/her infectious time span (so far) [0, *a*] infectees. When he/she has had age *c* ∊ [0, *a*], his/her infection rate was *β*(*c*). By now, the age of the focal individual/the infector is *a*. The probability that the infectee is now still infectious reads *κ*(*a* − c)). The rate of direct or indirect observation of the infectee with age of infection *a* − *c* is 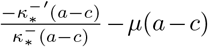. A detected individual triggers a successful tracing event with probability *η*_DCT_. Hence, the contribution to the removal rate of our focal individual due to tracing is given by

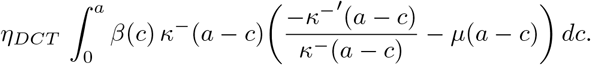

##### Proposition B.0.2

*The probability to be infectious for a symptomatic/asymptomatic individual at age of infection a for one-step contact tracing follows the following system of integro-differential equations*,

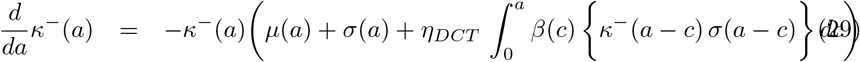

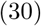

*with κ*^−^ (0) = 1.

##### Proof

The proof parallels that of proposition B.0.1; we only need to know that individuals have the rate of direct detection *σ*(*a* − c); this rate replaces the expression (detection rate for recursive tracing) 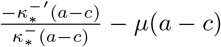

### Forward tracing

In contrast to backward tracing, in forward tracing the position of a focal individual in the tree/forest of infecteds matter. If I’m the primary infected person (generation 0), I have no infector (inside of the population). I cannot be traced by forward tracing. The first generation (infectee of generation 0) can only be traced via the primary infecetd person. The second generation can be traced by the zeroth and first generation, and so on.

That is, for forward tracing, the “generation” of an individual does influence the probability to be infectious at age of infection *a*. “Generation” does refer to generation of infection; the primary case has generation 0, those infected by the primary case have generation 1 etc. We denote by 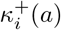 the probability to be infectious at age of infection *a* for an individual of generation *i* ∊ ℕ_0_ under forward tracing.

Furthermore, we introduce the probability to be infectious at age of infection *a* in case that we do not have contact tracing (*p* = 0) for symptomatic/asymptomatic infecteds,

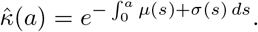

In order to determine 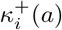, we first introduce and investigate

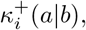

which is the probability that an individual in generation *i* is infectious at age of infection *a*, if we condition on the fact that the infector has age of infection *a* + *b* (s.t. the infector – at the time of the corresponding infectious contact – has has age of infection *b*). We find the following result

#### Proposition B.0.3

*In case of recursive tracing we have for i* > 0

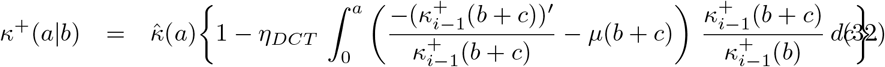

#### Proof

Our focal individual is infectious if it did not recover already without contact tracing, times the probability that no tracing event did remove the individual from the class of infecteds,

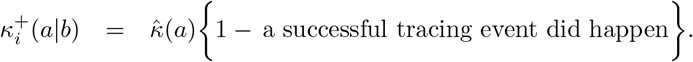

In order to obtain the probability for a successful tracing event, we first note that we know that the infector has been infectious at (his/her) age of infection *b*, s.t. the probability for him/her to be infectious at age of infection *b* + *c* reads

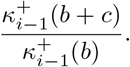

As before, for recursive tracing, the detection rate is the hazard rate minus the rate to recover spontaneously/unobserved,

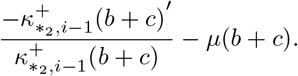

Hence, the desired probability reads a successful tracing event did happen 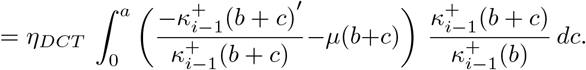 □

#### Proposition B.0.4

*In case of one-step-tracing we have*

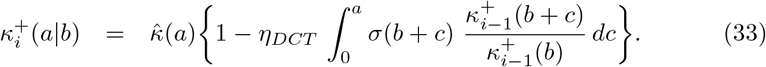

##### Proof

The argument parallels that of proposition B.0.3. We only need to take into account that in one-step tracing the infectee has to be detected directly, what happens at rate *σ*(.). This rate replaces the hazard rate minus the spontaneous recovery rate.

□

In order to determine the desired probability 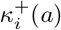 we remove the condition in 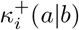. Thereto we determine the probability density for an infector to have age *b* of infection *b*. The net infection rate is *β_i_*(*b*) *κ_i_*_−1_*(b*). Therefore, the distribution of the age of the infector at the time of infection is given by+

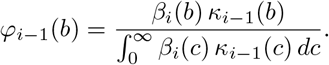

#### Corollary B.0.5

*In one-step tracing as well as in recursive tracing, we have for i* > 0

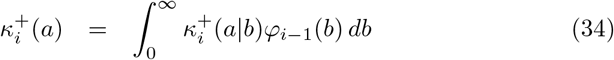

*where we have to use for* 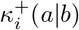 *the solution for one-step or recursive tracing, depending on the scenario chosen*.

We find an iterative formula. Analysis for *p* small as well as numerical analysis (for general *η_DCT_* ∊ [0,1]) shows that the convergence is rather fast: after 3-5 generations, the 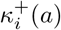 are basically converged.

### Full tracing

Full tracing is just a combination of forward- and backward tracing. Let *κ_i_*(*a*) denote the “survival probability” for a target individual of generation *i* under full tracing.

In order to find *κ*_0_(*a*), we only need to understand that the primary infected individual can only be traced by downstream infecteds, that is, is only exposed to backward tracing. Also the next generations have – without forward tracing – just the “survival” probability *κ*^−^(*a*). We need to multiply this probability with the probability not to be target of a forward tracing event in order to obtain *κ_i_*(*a*).

That is, we use the argument for forward tracing, where we replace 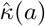 (survival probability without tracing) by *κ*^−^ (a) (survival probability under backward tracing only), and get immediately the following result (notation is an obvious extension of the notation above).

#### Proposition B.0.6

*In case of recursive tracing we have for i* > 0

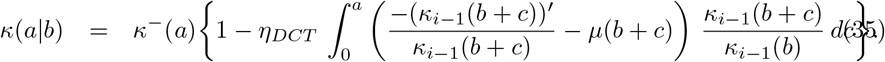

#### Proposition B.0.7

*In case of one-step-tracing we have*

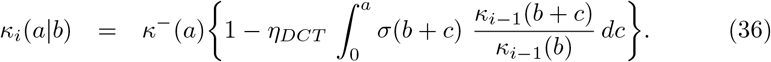

With

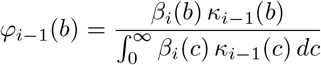

and for recursive contact tracing (one-step: parallel formula)

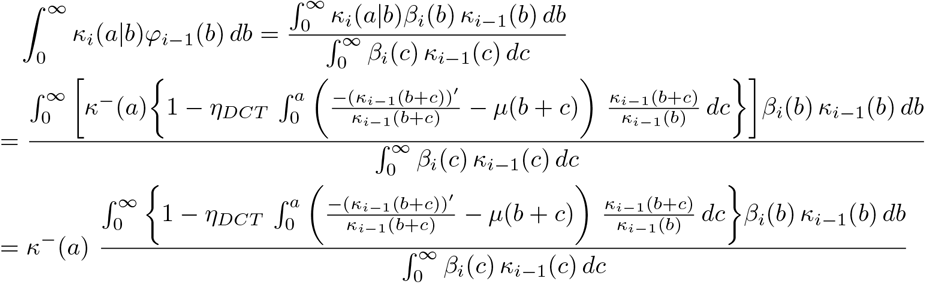

we finally get the result for full tracing (we summarize all necessary equations):

#### Theorem B.0.8

[recursive tracing] *Let*

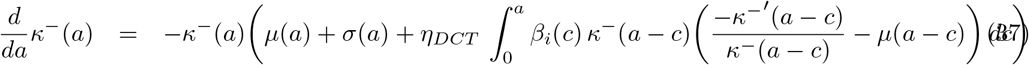

*with κ*^−^ (0) = 1. *Then*,

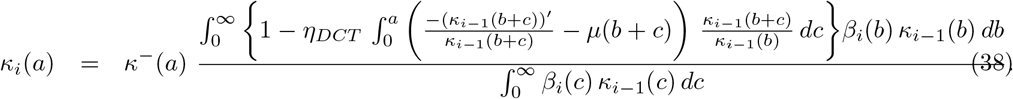

#### Theorem B.0.9

[one-step tracing] *Let*

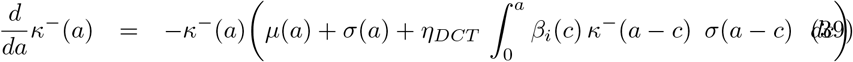

*with κ*^−^(0) = 1. *Then*,

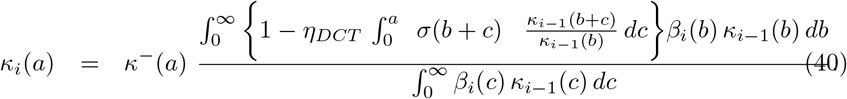

Note that it is straightforward (but tedious) to simplify these equations, s.t. they become handy for the numerical analysis.

### Reproduction number and exponential growth

A direct consequence of the dynamic, age-structured model is the possibility to determine the reproduction number and the exponential growth rate right away from the survival probability *κ_∞_*(*a*). The effective reproduction number is simply given by

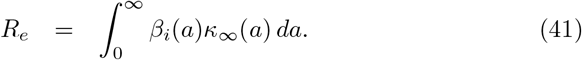

The exponent *λ* of the exponential growth in the onset *S*(*t*) ≈ *S*_0_ ≈ *N* can be determined by the unique real root of the equation

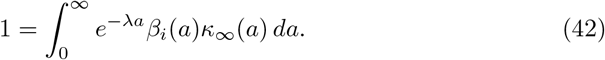

From here, we obtain *T*_2_ = ln(2)/*λ*.

### Reproduction number in case of DCT

Recall that we consider a homogeneously mixing population, where a fraction *p_app_* of individuals have a DCT device. Only contacts between these individuals can be traced, with probability *η_DCT_*. Let *R_eff_*(*p*) denote the reproduction number of a homogeneous model, where each contact is traced with probability *p*. Then, a straightforward generalization of the considerations above yields that non-app-users have the reproduction number *R_eff_*(0), while those with app have the reproduction number

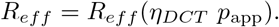

Since we assume homogeneous mixing, we obtain the overall reproduction number as

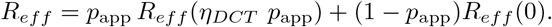

This formula is exact, but due to the nonlinearity in *R_eff_* (.) it cannot be simplified. However, if *p_app_*, *η_DCT_* ≪ 1, we can linearize at *p_app_* = 0, and find

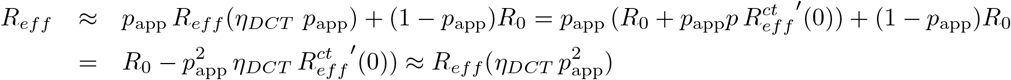

In lowest order, tracing within a subgroup of relative size *p_app_* and a tracing probability *η_DCT_* is equivalent with tracing the total population with a tracing probability 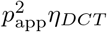.

## Results

We identify the parameters of the models as described below. Using numerical analysis, it is possible to estimate the influence of control measures on the spread of the infection. Particularly, we are interested in the effect of contact tracing and social distancing on the effective reproduction number and the doubling time. Moreover, the sensitivity of the results on parameters for which there is little data is explored. Here, our focus is the fraction of asymptomatic cases.

If we inspect Fig. 16, we find first of all that recursive tracing is more efficient than one-step tracing (one must not confuse one-step tracing with level-1 tracing: here, we only follow infectious contacts for one step). In practice, the tracing delay will lead to a situation between one-step tracing and recursive tracing: even if recursive tracing is the aim, as we loose time in tracing each contacts, after a few steps persons might already be recovered when traced. In any case, small tracing probabilities have only a minor impact. If we increase the tracing probability to 0.7-0.8, we find that an eradication of the infection is possible. On the other side, social distancing is most effective for small tracing probabilities. The combination of contact tracing and social distancing is most likely best in the middle range: a reduction of contacts by 30%, and a tracing probability around 0.6-0.7 will be able to reduce the doubling time considerably, and even bring *R_e_* down to values around 1.

The parameter values are not all precisely known. In particular, at the present time, the fraction of undiagnosed cases is rather unclear. In the literature, there are numbers between 1.2% and 95%. The parameter scan in Fig. 17 indicates that the results are rather stable for a wide range of *α* (from 10-50%). A major change can be observed above 70%. This figure might imply that the results are rather stable against the parameter choice in *α*.

### Choice of parameter functions

The medical investigations yield particularly data on:

- Incubation period (time to the onset of symptoms/diagnosis)
- latent period (time until infectivity becomes positive) and viral load (a proxy for infectivity)
- fraction of asymptomatic cases.

#### Incubation period

Time to symptoms: We use a Γ distribution, density

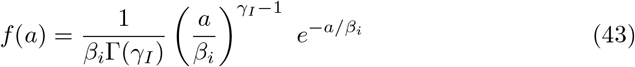

with parameters *β_i_* = 2.44, *γ_I_* = 3.06 days. Let 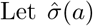 be the conditioned detection rate: It is only valid under the condition that an individual is detected, indeed. That rate is just the hazard rate of *f*(*a*),

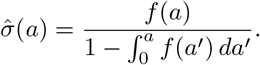

We later need to compute the rate *σ*(*a*) for the model, where we take into account the fraction of asymptomatic persons.

#### Infectivity

Given that the onset of symptoms of a person at age *A*_0_ (a Gamma-distributed random variable, as described above), the infectious period starts *A*_0_ − Δ*_onset_*, where Δ*_onset_* is a fixed time span (we choose Δ*_onset_* = 3 days). If *a*_0_ − Δ < 0, then the infectivity period starts right away at the time of the infection. That is, max{Δ_0_ — Δ*_onset_*, 0} is the latent period. We furthermore assume that the infectious period is a fixed (deterministic) time span *T*_con_.

This assumption about the latent period is an input for the age-dependent infection rate as well as for the recovery rate.

#### (b.1) Recovery rate

If *A*_0_ ~ Gamma(*β_i_*, *μ_I_*) is a random variable that states the onset of symptoms, we aim at the distribution of the recovery age *A_r_* = max{*A*_0_ − Δ*_onset_*, 0} + *T*_con_. Clearly, *P(A_r_* < *a*) = 0 for *a* ≤ *T*_con_. A short computation yields for *a* > *T*_con_

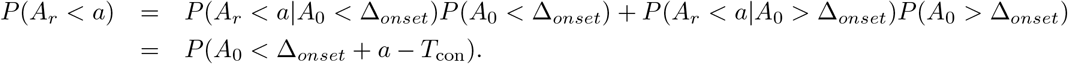

Hence, in total we have

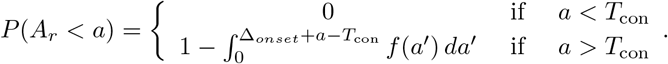

As the cumulative distribution involves a jump, the hazard rate (recovery rate) incorporates a delta peak. However, for the practical implication, we replace the jump by a steep linear increase during a small age interval. The corresponding hazard rate yields the removal rate 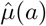, conditioned on the fact that a person will not be diagnosed by recover spontaneously. The parameter *T*_con_ is computed below.

#### (b.2) Infectivity

We again denote by *A*_0_ the random variable that states the onset of the symptoms, *A*_0_ ~ gamma(*β_i_*,*γ_I_*).

The ingredient for the infectivity is an approximation of the virus load, given by a shifted Gamma distribution,

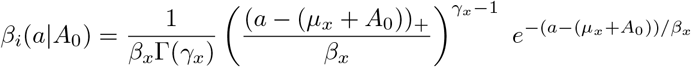

with the understanding that this formula is only valid for *A*_0_ > − *μ_x_*. Here, (*x*)_+_ = 0 for *x* < 0 and (*x*)_+_ = *x* if *x* > 0. The parameters are given by:

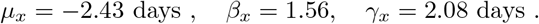

If *A*_0_ + *μ_x_* < 0, we assume that symptoms start right away. Hence,

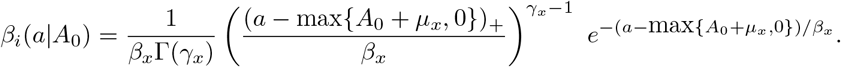

Then (as in our case *μ_x_* < 0). *β_i_* is proportional to

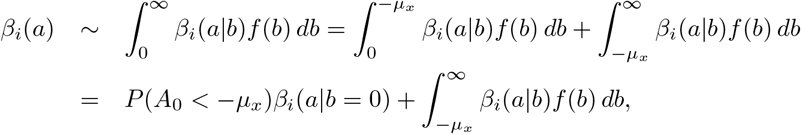

where *f*(*b*) is given in (43). The unknown proportionality constant models the number of contacts per day. This constant is calibrated s.t. the basic reproduction number *R*_0_ is 2.5.

We define *T*_con_ by

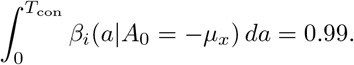

### Asymptomatic cases

We define the effective detection rate *σ*(a) and the effective removal rate *μ*(*a*), based on the the conditioned rates 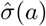 and 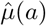, and the fraction of asymptomatic cases α. The probability to be infectious is a convex combination of the probabilities to be infectious for a symptomatic resp. asymptomatic individual (where we understand that an “asymptomatic individual” is not only asymptomatic at a given time, but will stay undiagnosed until recovery). That is,

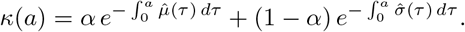

In order to find the effective rates, we compute the hazard rate of *κ*(*a*),

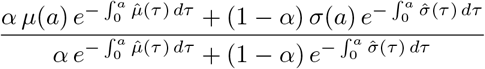

From here, we immediately find the desired definitions for *μ*(*a*) and *σ*(*a*),

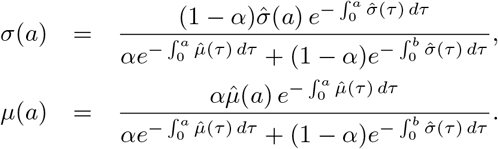

## C Model benchmarking

Monte Carlo models rely on computer code, and therefore must be verified to work as designed. We do this in two ways: a) by comparing results to deterministic models, and b) by comparing the results from two implementations written independently by two teams using two different programming languages and toolsets. One example for (a) is shown in Fig. 18. The deterministic models cannot model all effects we consider, therefore POC testing and social distancing was turned off in the Monte Carlo models, and other parameters were chosen as summarized in Tab. 3. The parameters in the ODE model do not have straight-forward correspondence to the parameters in the table, so they were tuned such that the outcome matches the other models as well as possible.

The analytic models are, strictly speaking, not valid once a sizable fraction of the population has been exposed, since they do not model non-linear effects such as two exposed people meeting each other. Nevertheless we show the full outbreak curves here.

**Table 3:** Table of settings used to benchmark the four models against each other.

| Parameter/Setting | Benchmark value |
| --- | --- |
| Disease and population |  |
| Size of population | 10k |
| Population structure | uniform |
| Transmission probability ($\beta_i$) | $\begin{pmatrix} 0.0595 \\ 5.0 \\ 3.2 \end{pmatrix}$ |
| Contact rate |  |
| R0 |  |
| Trans. prob. curve ($\mu, \gamma, \beta$) | (-2.42, 2.08, 1.56) |
| Incubation time curve ($\mu, \gamma, \beta$) | (0, 3.06, 2.44) |
| Fraction symptomatic ($\alpha$) | 0.5 |
| Asymptomatic trans. scaling ($\eta_{as}$) | 1.0 |
| Interventions |  |
| Interventions start ($f_i$) | 0.00 |
| Quarantine duration | 14 days |
| Tracing |  |
| Reported from symptoms ($f_m$) | 1.0 |
| Trace back ($\Delta T_{trace}$) | 14 [days] |
| App coverage ($p_{app}$) | 1 |
| Tracing efficiency ($\eta_{DCT}$) | 0.7 |
| Tracing order | 1 |
| Trace uninfected contacts | False |
| Tracing delay ($T_{delay}$) | 0 [days] |

The ODE model matches quite well with the Python and C++ models, but it has the advantage of having been tuned to achieve as good a match as possible. The age-structure model has a time offset compared to the others, but the time when the outbreak takes off in the IB models is very variable so such offsets are not relevant.

Even though both individual-based models implement the same disease parameters and intervention protocols, there are a number of implementations details that are treated differently and can lead to slightly different outcomes. Examples of this are whether nor not people with a 0 days latency period can infect others on the same day they themselves became infected, or whether we allow someone to still infect others on the day they become symptomatic and are quarantined.

Figs. 19 and 20 compare what happens for two implementations of a latency period of zero days. In one implementation, it is possible for the carrier to infect others on the same day he or she became exposed. In the other implementation, a carrier can infect others only on days after he or she became exposed.

**Figure 19:**
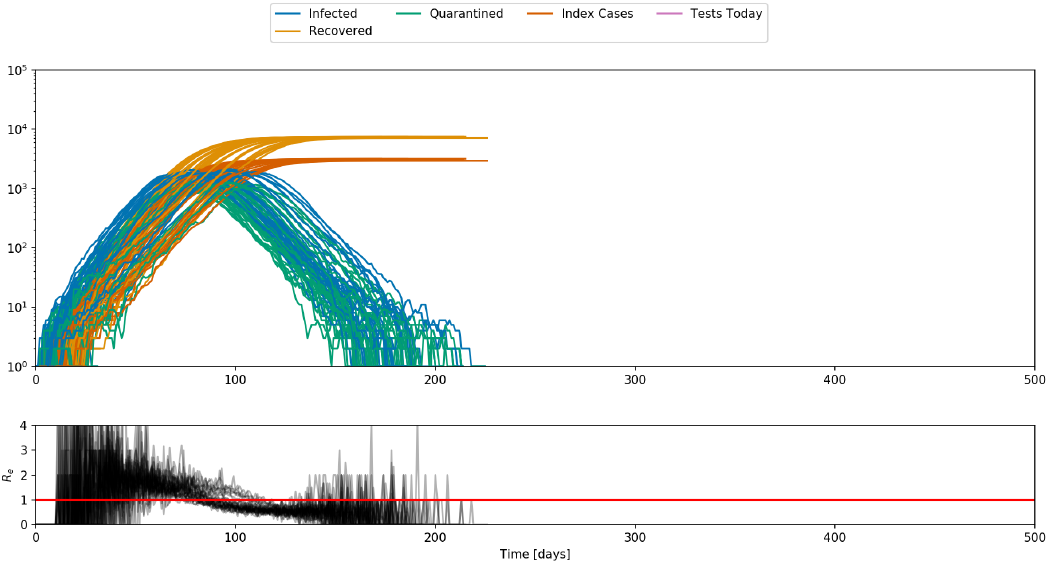
Outbreak dynamics when someone with zero days latency period cannot infect others on the same day he or she became infected.

**Figure 20:**
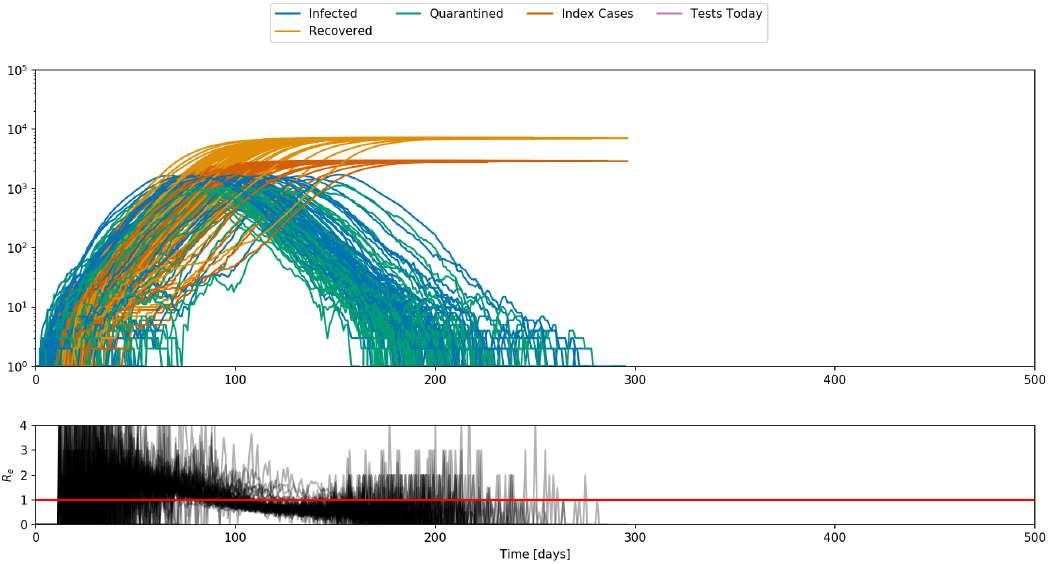
Outbreak dynamics when someone with zero days latency period can infect others on the day he or she became infected.

One big difference between the IB models is that the Python model accounts for the case where a contact is traced from an index case who did not infect the contact, but in the time between when the contact took place and when the person was traced, the contact was infected by someone else. In the C++ model, this special case is not considered. The effect on the outcomes is small, especially when taking a look-back time of only 7 days as done for most of the main scan.

Fig. 21 shows how the population structure affects the course of the epidemic. Social graph populations lead to long-term changes in the outbreak dynamic that are not accurately captured by just the reproductive number reached after interventions.

**Figure 21:**
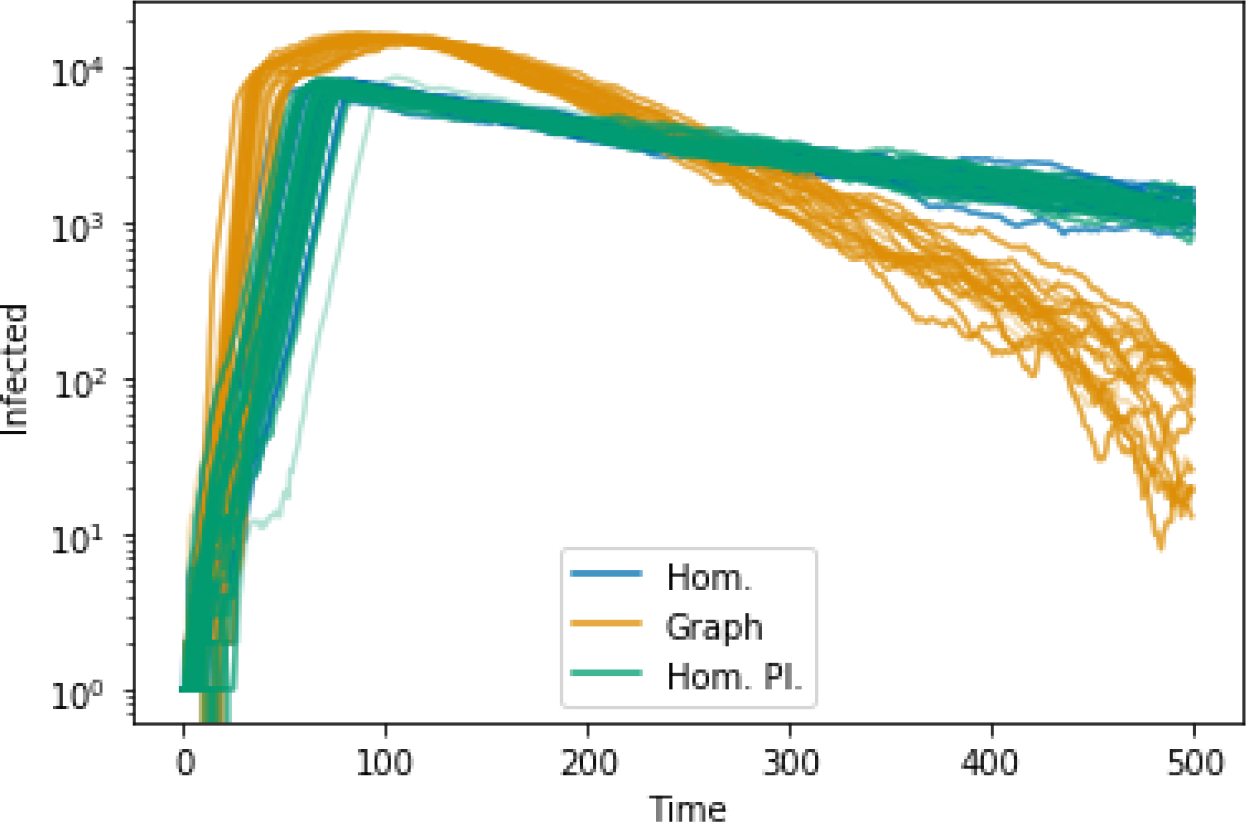
The number if infectious people each day for a social graph and a homogeneous population is compared.

Fig. 22 is similar to Fig. 6 but for different parameters. In this scenario, the reproductive number changes both due to the interventions and naturally because a large enough fraction of the population becomes exposed early on that non-linear effects are important.

**Figure 22:**
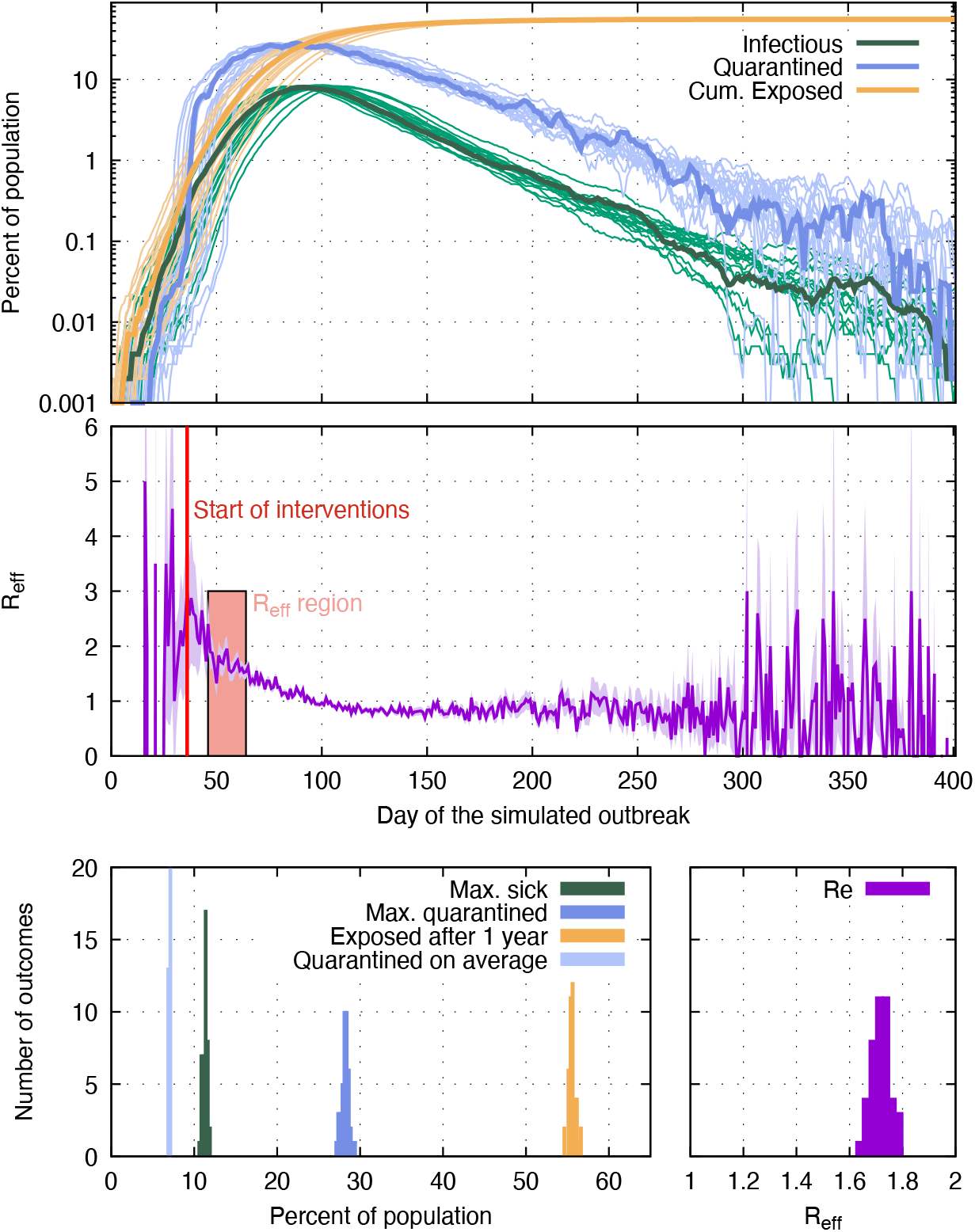
Same as Fig. 6 changing the following parameters: *p_app_* =0.6, *α* ·*f_m_* =0.6.

## D Results from all scenarios

The results for the main parameter scan are shown in Figs. 23 through 28. Results for the scan with different timing and delays are shown in Figs. 29 through 34

**Figure 23:**
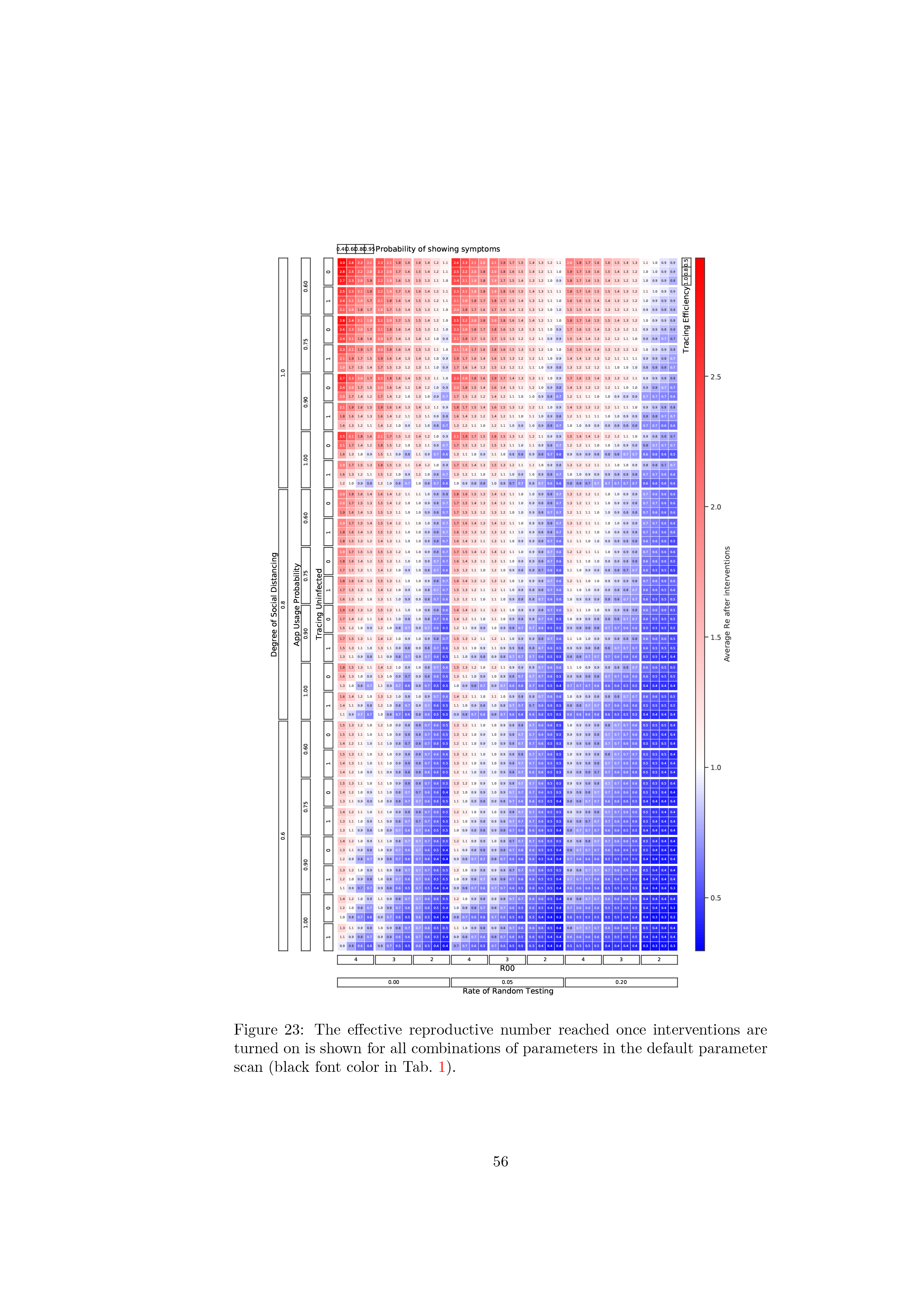
The effective reproductive number reached once interventions are turned on is shown for all combinations of parameters in the default parameter scan (black font color in Tab. 1).

**Figure 24:**
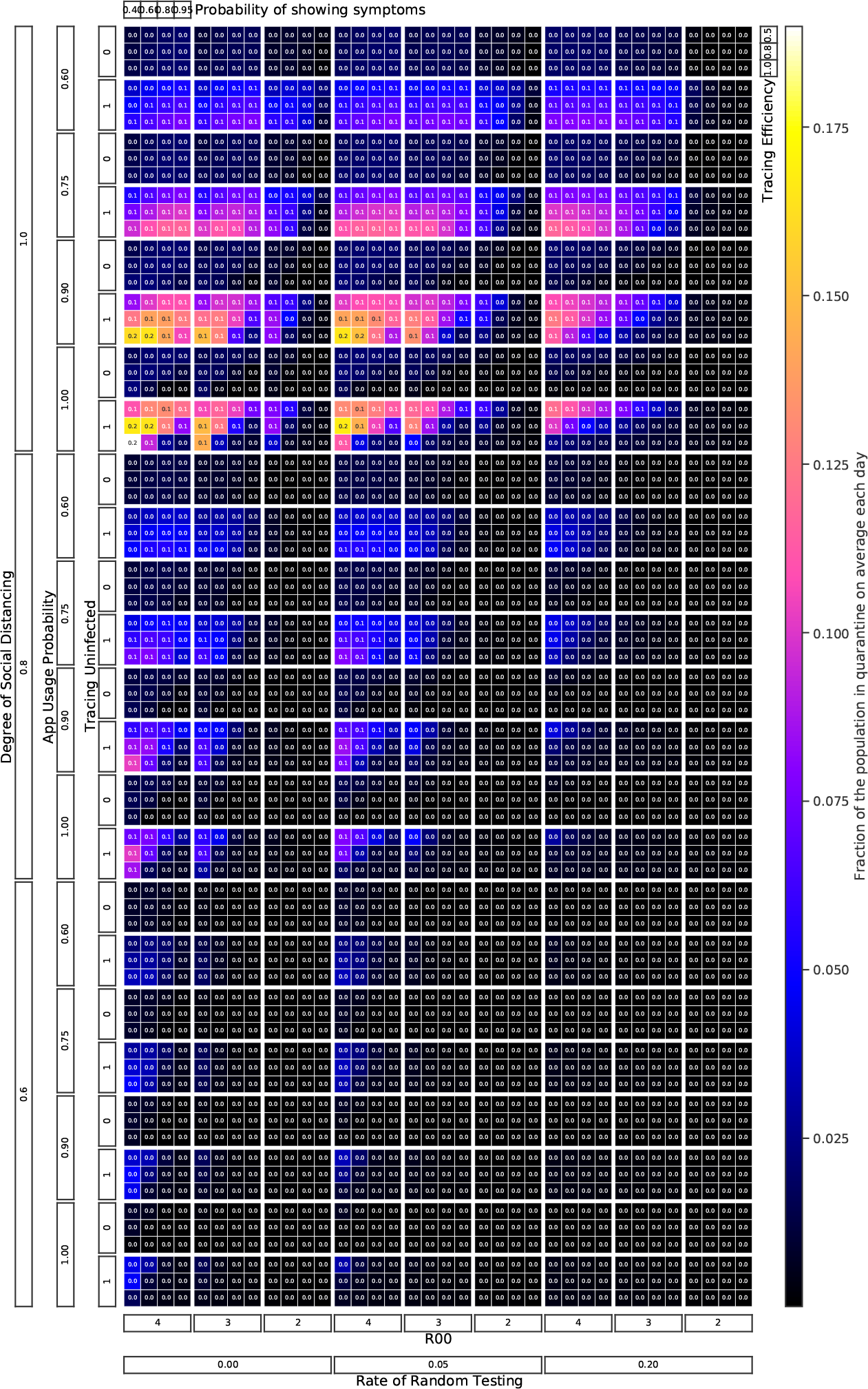
The fraction of the population quarantined on average over one year of the outbreak is shown for all combinations of parameters in the default parameter scan (black font color in Tab. 1).

**Figure 25:**
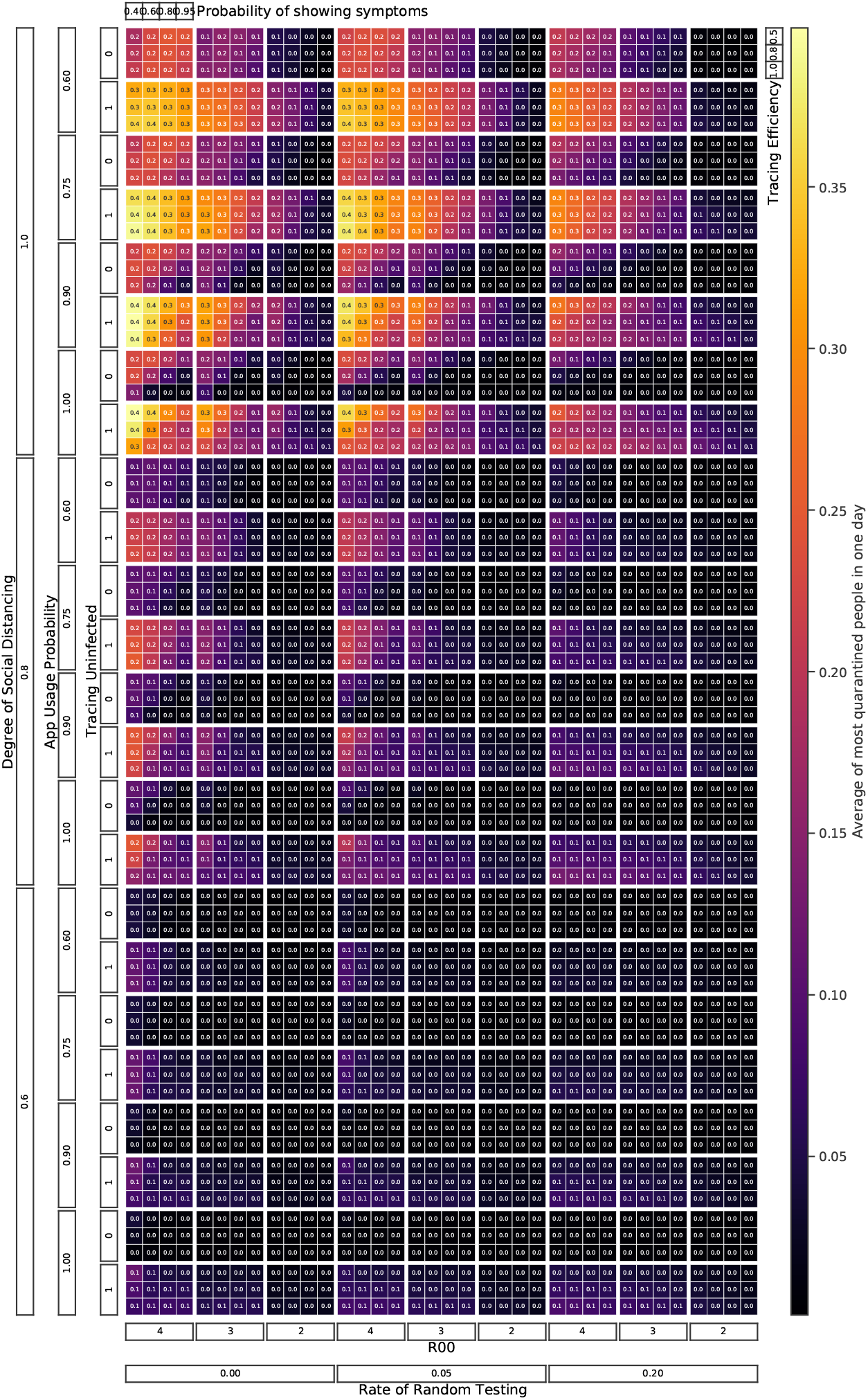
The fraction of the population that is quarantined on the day when most people are quarantined is shown for all combinations of parameters in the default parameter scan (black font color in Tab. 1).

**Figure 26:**
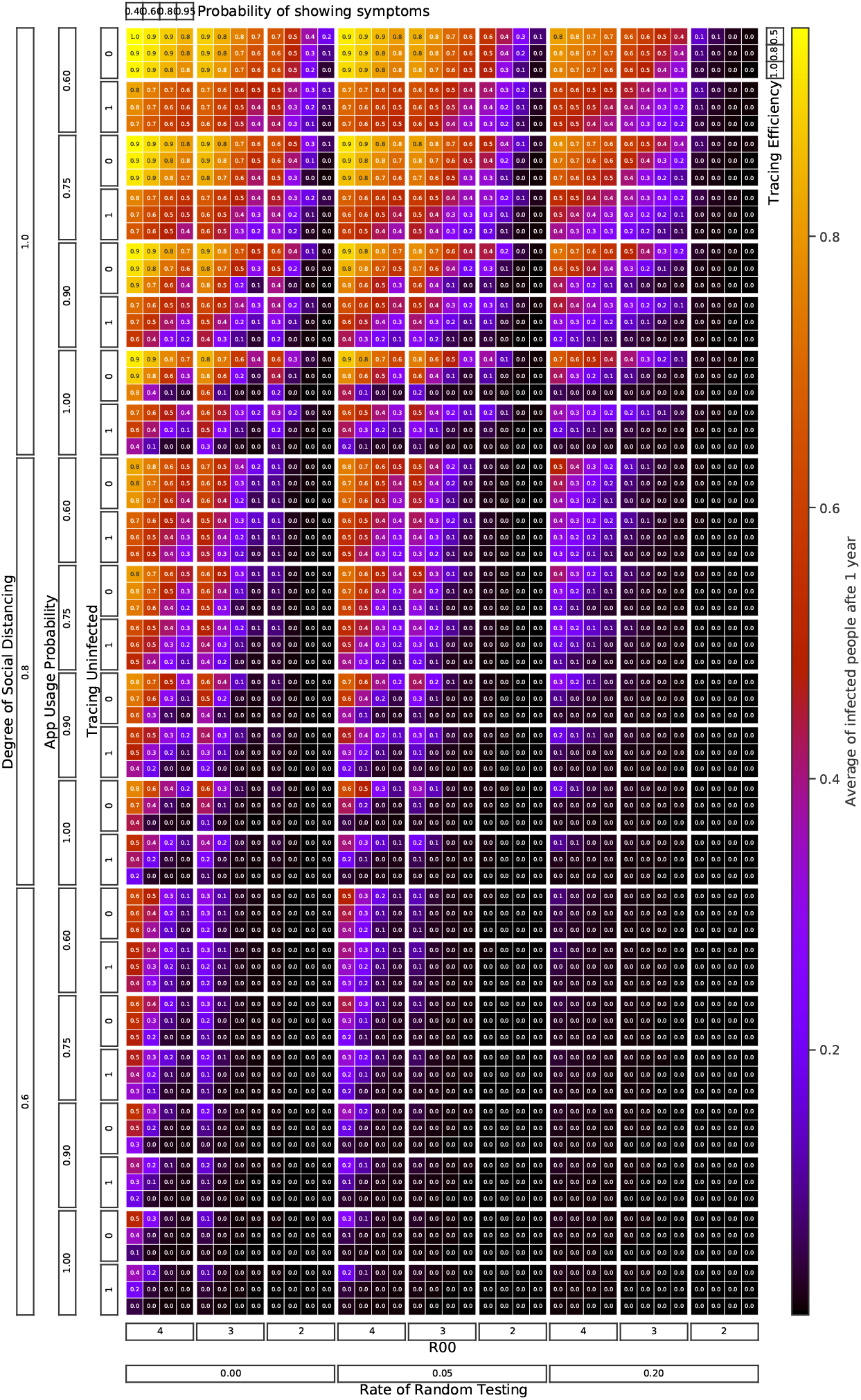
The total fraction of the population that has been exposed one year into the outbreak is shown for all combinations of parameters in the default parameter scan (black font color in Tab. 1).

**Figure 27:**
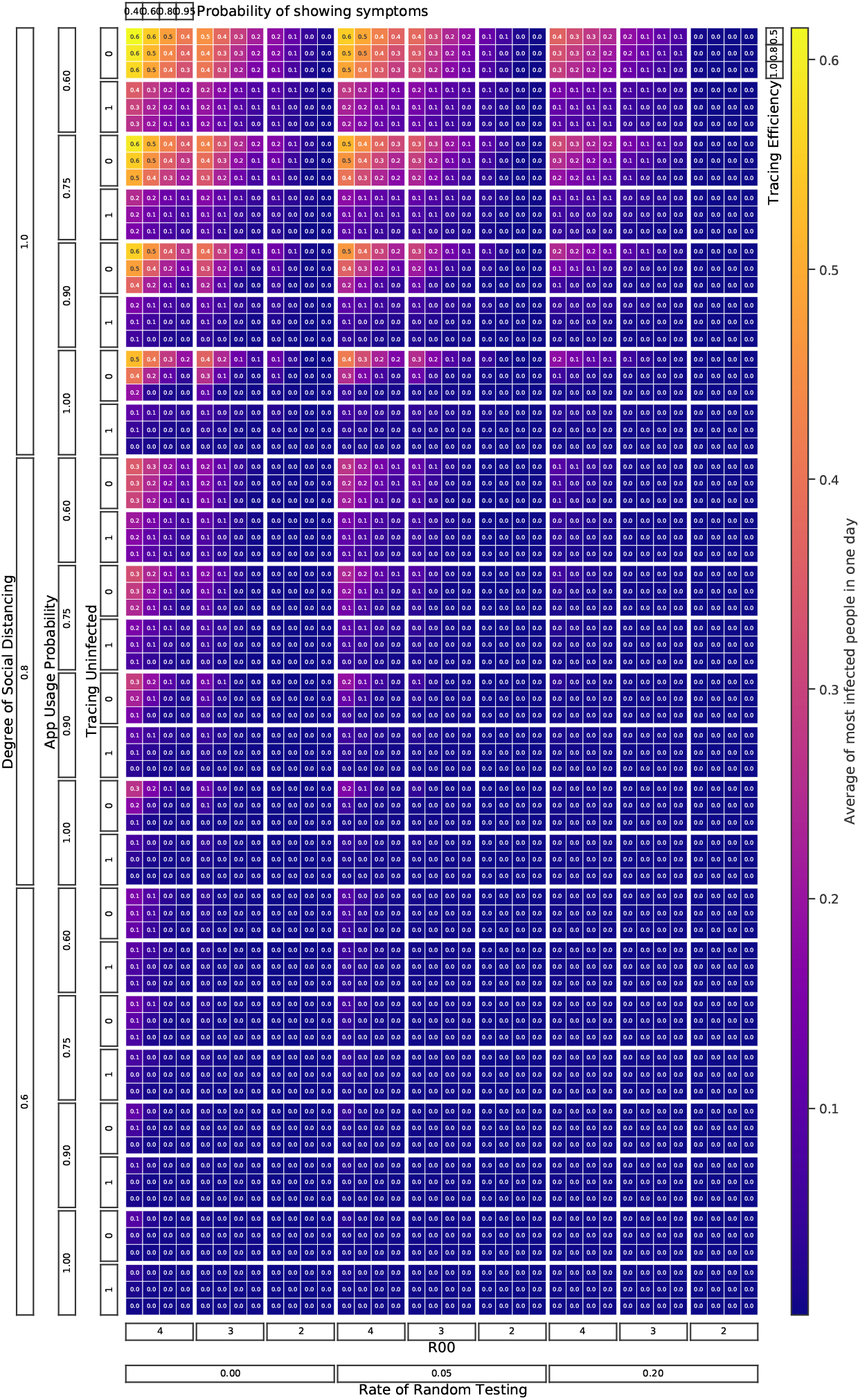
The fraction of the population that is sick on the day when most people are sick is shown for all combinations of parameters in the default parameter scan (black font color in Tab. 1).

**Figure 28:**
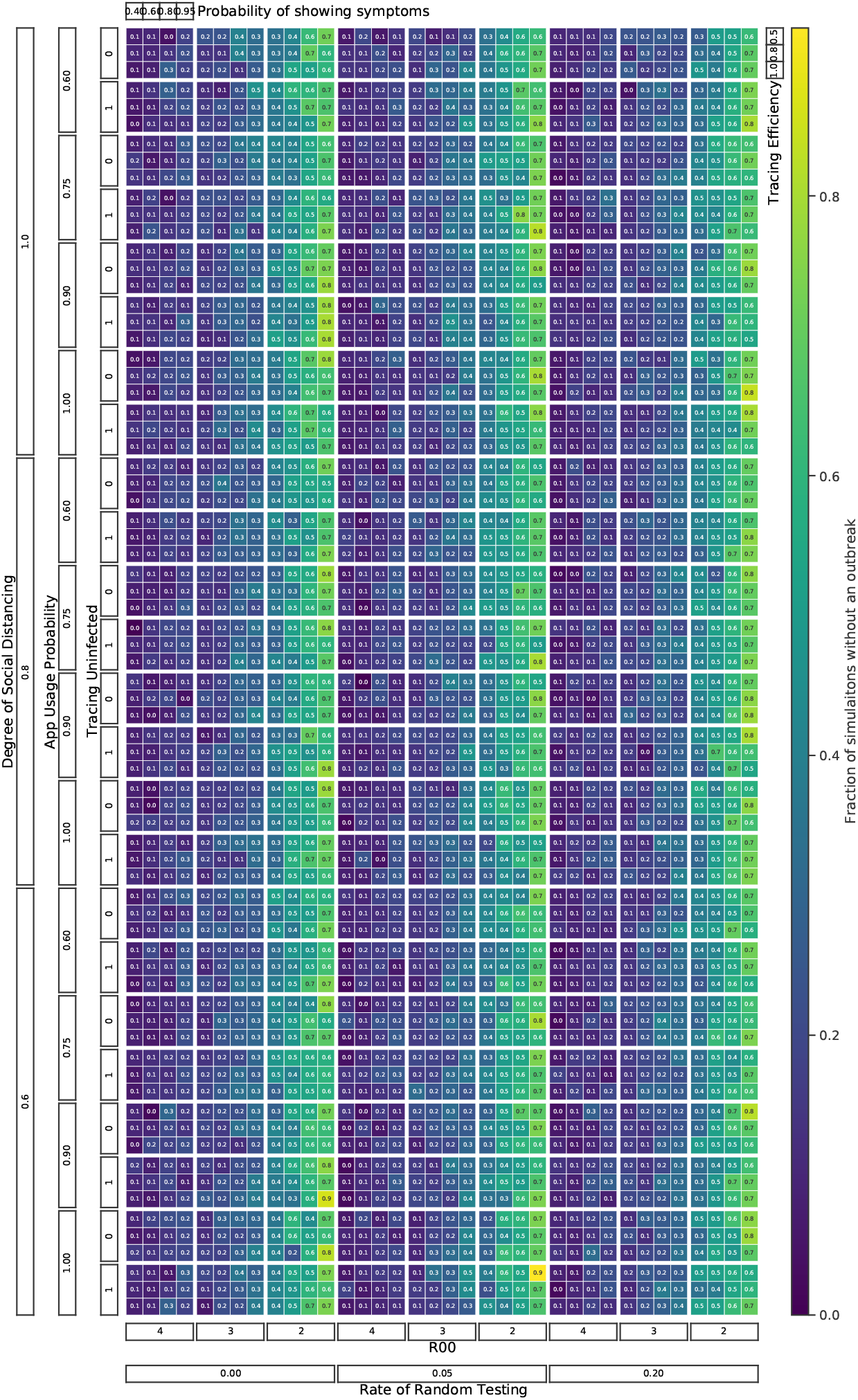
The fraction of MC runs where the outbreak stopped (by chance) before the threshold number of infected people to start interventions was reached.

**Figure 29:**
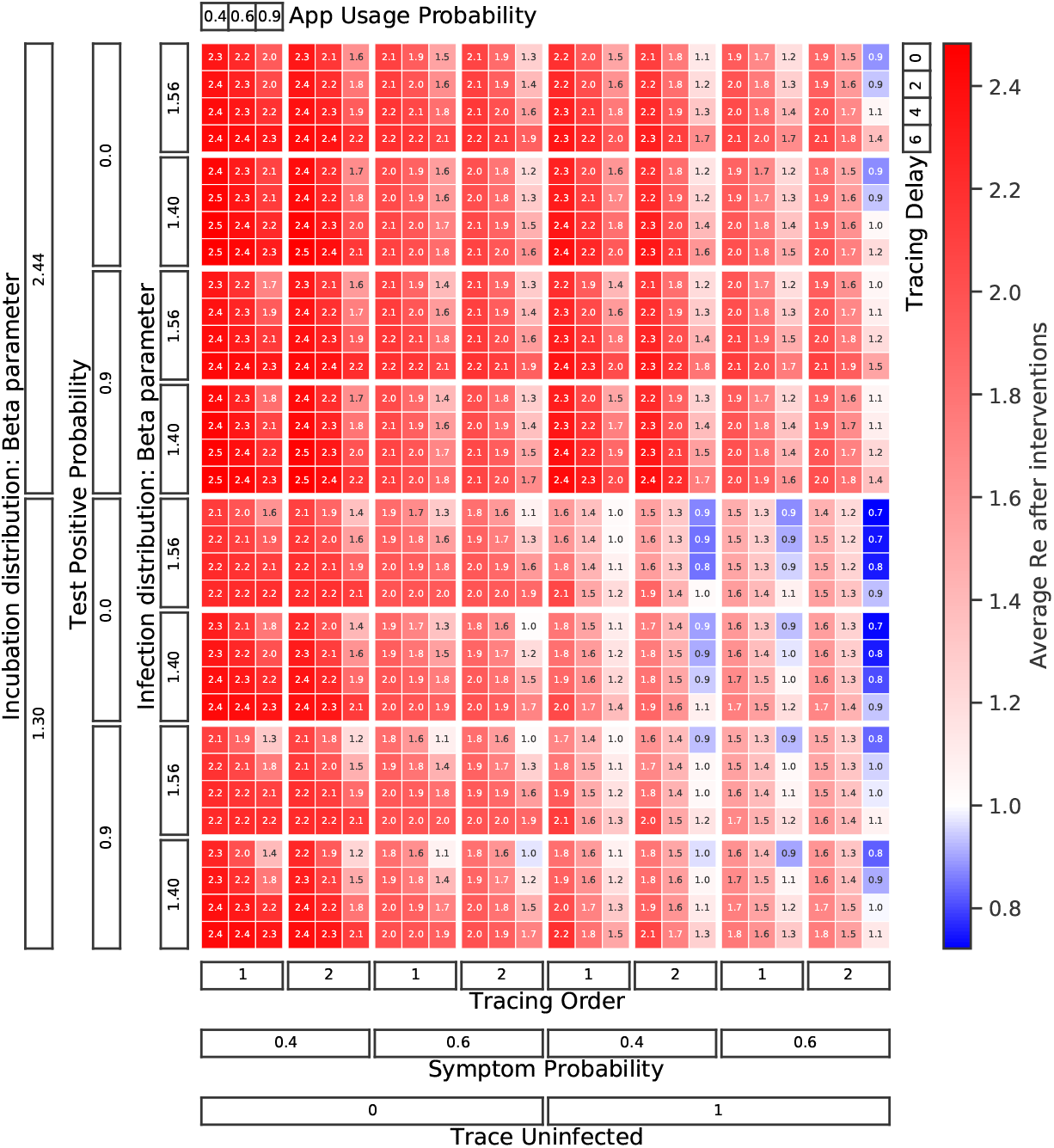
The effective reproductive number reached once interventions are turned on is shown for all combinations of parameters in the timing and delay parameter scan

**Figure 30:**
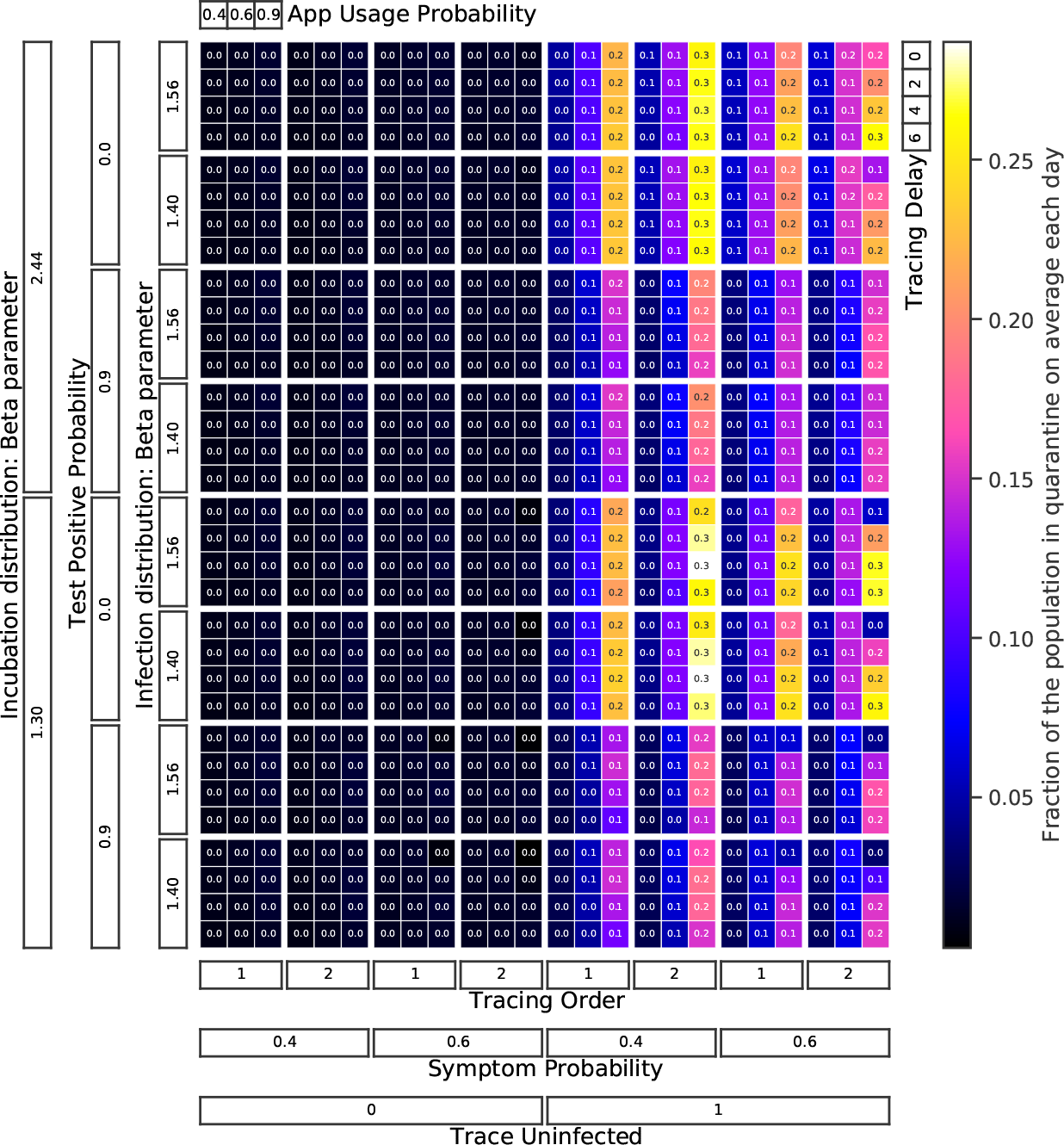
The fraction of the population quarantined on average over one year of the outbreak is shown for all combinations of parameters in the timing and delay parameter scan.

**Figure 31:**
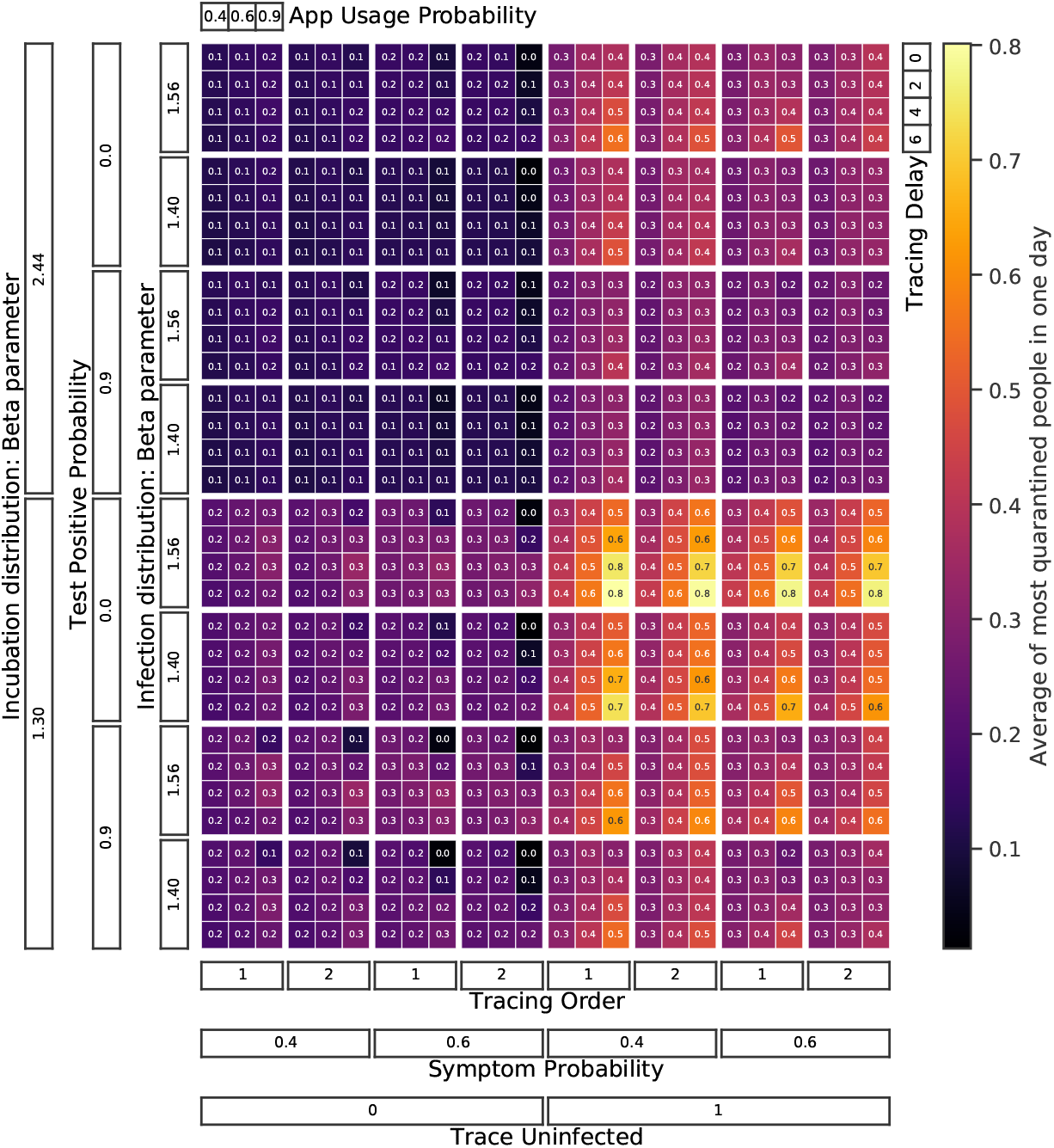
The fraction of the population that is quarantined on the day when most people are quarantined is shown for all combinations of parameters in the timing and delay parameter scan.

**Figure 32:**
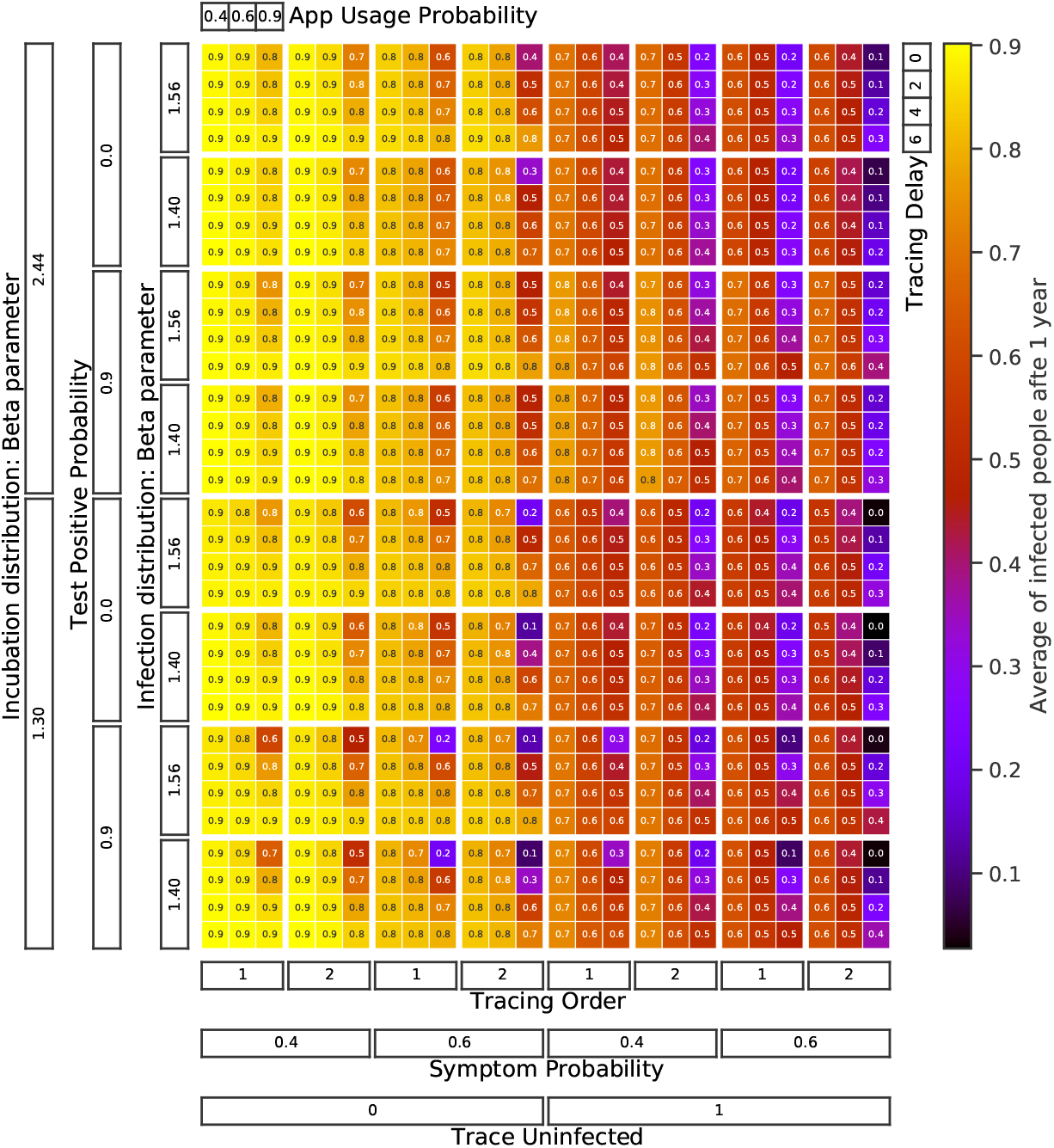
The total fraction of the population that has been exposed one year into the outbreak is shown for all combinations of parameters in the timing and delay parameter scan.

**Figure 33:**
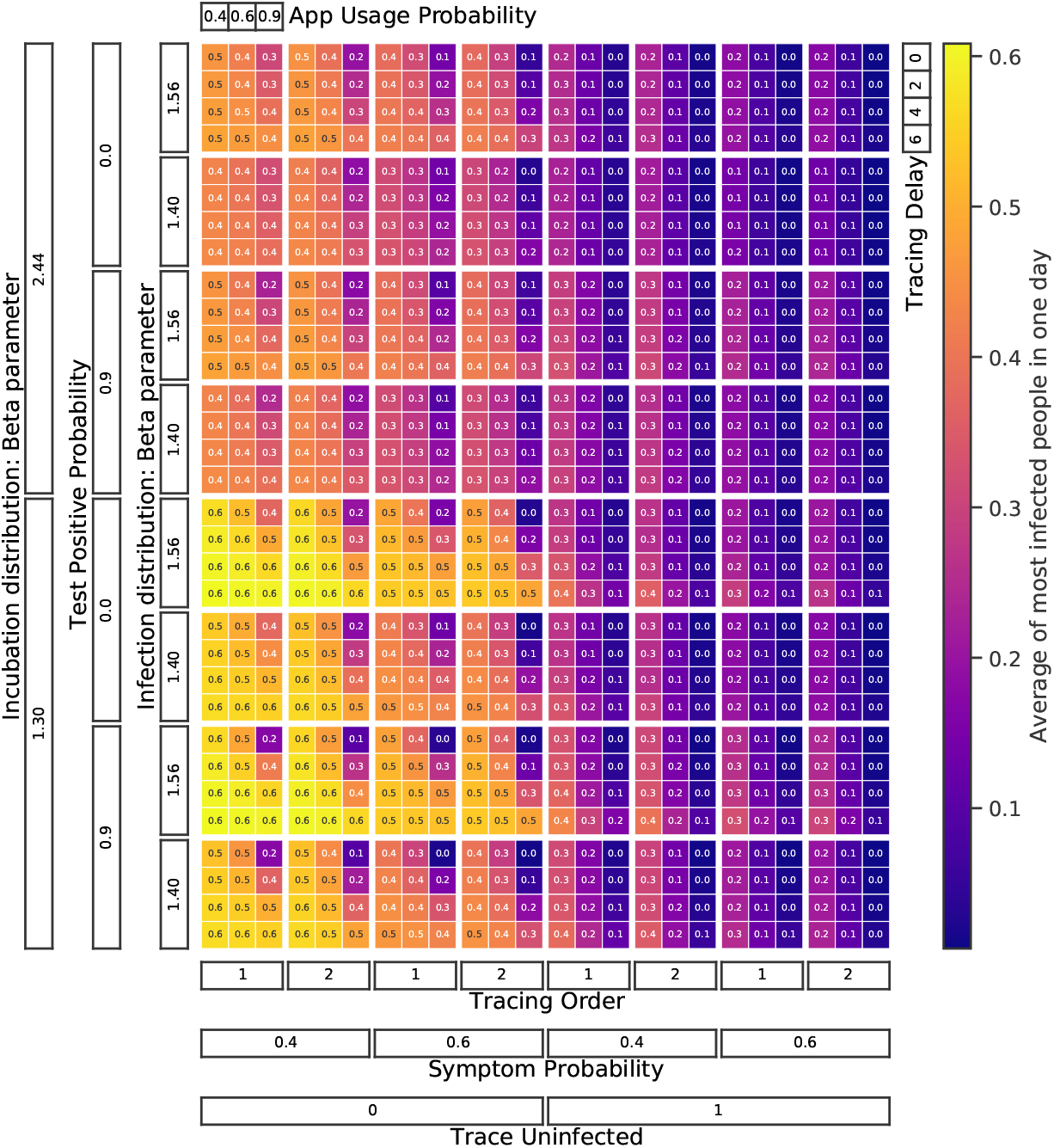
The fraction of the population that is sick on the day when most people are sick is shown for all combinations of parameter in the timing and delay parameter scan.

**Figure 34:**
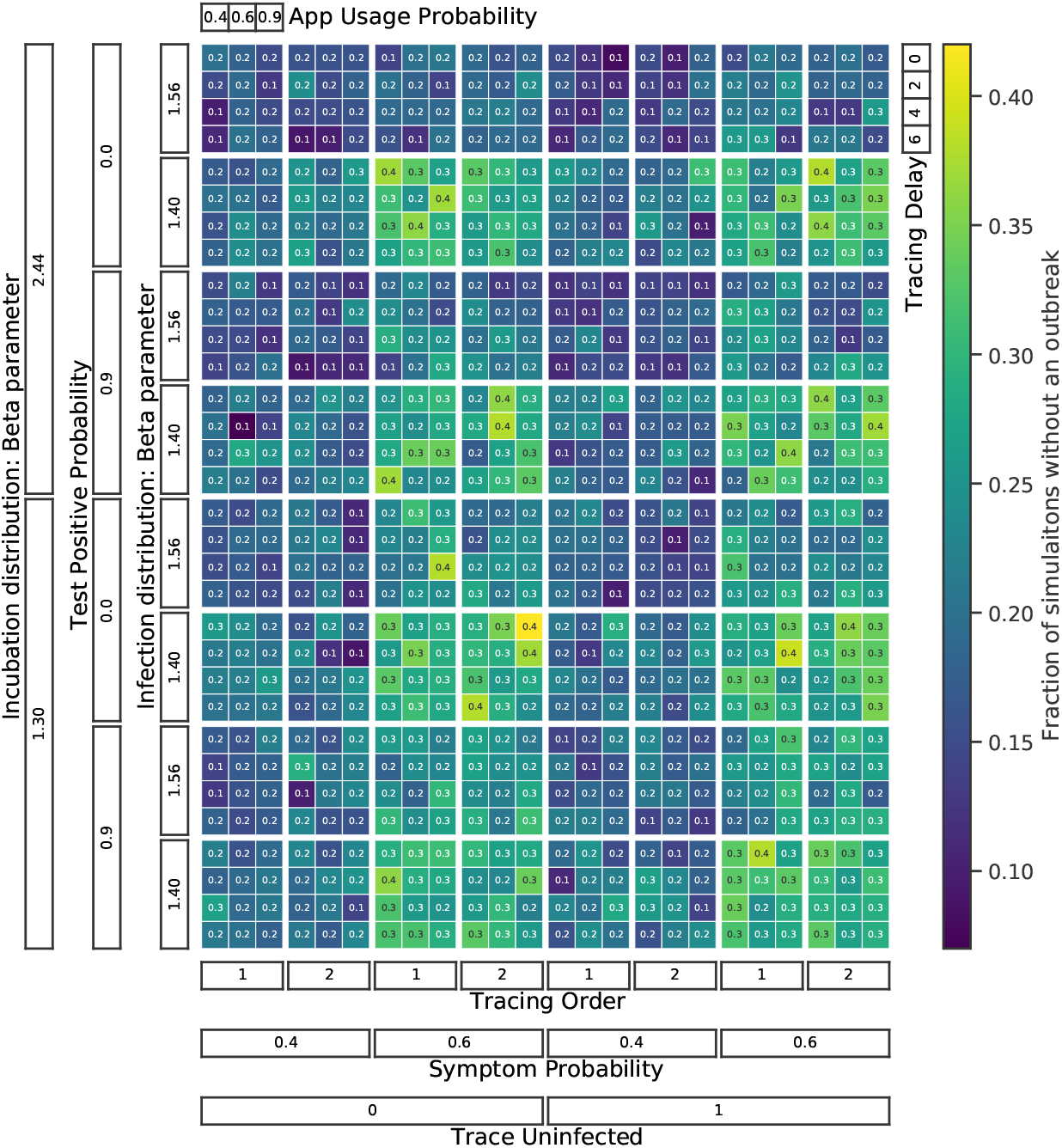
The fraction of MC runs where the outbreak stopped (by chance) before the threshold number of infected people to start interventions was reached (timing and delay parameter scan).

## Footnotes

1 A similar second infectious state has been implemented by Ref.[29] to describe individuals that can be tested positive beyond their infectious period. While the viral load in this second state is still large enough for detection, it is too small for further infection.

## Notes

### Competing Interest Statement

The authors have declared no competing interest.

### Funding Statement

No external funding was received for this work. The Computational Center for Particle and Astrophysics (C2PAP) and the Max Planck Institute for Physics kindly allowed us to make use of their computing resources.

### Author Declarations

No approval was necessary for this work. It is purely a simulation study.

